# A Systematic Review of Datasets to Research the Effect of Policy and Place on Immigrants’ Health

**DOI:** 10.1101/2022.01.30.22270115

**Authors:** Chenoa D. Allen

## Abstract

State and local immigrant policies—which regulate immigrants’ rights and access to services and material resources—shape self-rated health and health care access for Hispanic/Latino immigrants. However, due to data limitations, we know little about their impacts on non-Hispanic immigrants or on objective measures of health. To help researchers address these gaps, this systematic review creates a comprehensive resource indexing nationally-representative US datasets that can be used to measure the effects of policy and place on immigrant health. Following PRISMA guidelines, I searched ICPSR and federal data repositories for nationally-representative, population-based, longitudinal or repeated cross-sectional datasets that included health/health care access, race/ethnicity, and state or county of residence. I extracted information about race, ethnicity, migration, language, and geocodes in each dataset for each year, 2005–2021. Data searches (final search December 2021) identified 54 datasets, many of which have never been used to study the effects of policies on immigrant health. These data sources have unexploited potential to advance this literature. For example, longitudinal data sources will allow researchers to examine the effects of policies on the same individuals over time, while datasets with detailed measures of race/ethnicity and migration can be used to distinguish which immigrants are most impacted.

## INTRO

Nearly 46 million immigrants live in the United States, including 20.7 million naturalized citizens, 14.5 million legally-present noncitizens, and 10.5 million undocumented immigrants (1). In addition, 16.3 million US-born children have an immigrant parent (2). Immigrant health and well-being are shaped by demographic, economic, social, political, and environmental characteristics of the communities where they live (3, 4). There is growing recognition that federal, state, and local policies that increase immigration enforcement (the arrest and deportation of immigrants) are associated with worse health (4–6), reduced health care utilization (7, 8), less trust in government health agencies (9), and greater poverty (10) for immigrants and their children. In contrast, policies that expand immigrants’ access to employment, education, public benefits, and driver’s licenses are associated with better health (11–13) and health care access (14, 15).

These policies, collectively referred to as immigrant policies, may affect immigrants from all countries of origin, as well as their US-born co-ethnics. They may also shape multiple domains of health and mortality across the lifecourse (3, 16). However, current research is limited in scope with respect to the *health outcomes* and *immigrant populations* studied.

Most population-level research focuses on health care access (7, 8, 17) and single-item measures of self-rated health (11, 12). Because few studies include objective measures of morbidity and mortality (for exceptions, see 4, 5, 13), we do not yet know to what extent immigrant policies have measurable impacts on physical or mental health.

There is also limited evidence about which immigrants are most affected. Most studies examine effects on Hispanic/Latino immigrants; few studies evaluate the health impacts for the 21.4 million immigrants from Africa, Asia, Europe, and Oceania (18). Two studies that compare effects on immigrants of different races/ethnicities find that state immigrant policies affect Latino, Asian, and Black immigrants differently (17, 19). In addition, policy effects are expected to differ by legal status because state and local policies often target undocumented immigrants (3). However, few studies include direct measures of immigrants’ legal status (e.g., legal permanent resident, Temporary Protected Status (TPS), Deferred Action for Childhood Arrivals (DACA), undocumented/unauthorized (20)). By aggregating immigrants of different legal statuses, studies likely underestimate policy effects for undocumented immigrants and overestimate effects for legally-present immigrants. Some studies use country of birth and level of education to proxy for legal status (10), but these proxies are less accurate than direct measurements of legal status (21). Cross-survey multiple imputation methods can *impute* legal status in nationally-representative datasets. Under the right conditions, these methods can produce unbiased estimates of undocumented immigrants’ well-being, but are not suitable for all datasets and are not widely used (22).

These gaps in our understanding of policy effects are due in part to data limitations. Most research relies on three datasets: the American Community Survey (ACS) (10, 17), US birth records (5, 14), and the National Health Interview Survey (NHIS) (6, 12). These datasets are commonly used because they are among the most reliable and accurate sources of data about health and health insurance coverage (23, 24); they provide annual, repeated cross-sectional data so that researchers can examine changes in population health before vs. after policy implementation; and they include geocodes, which are necessary for researchers to estimate the health effects of state and local policies. However, each has limitations. The ACS and birth records have large sample sizes that allow disaggregation by race/ethnicity and country of birth, but include few measures of health/health care access. NHIS has more detailed health data, but the utility of NHIS for studying immigrant families is limited after the 2019 redesign, due to smaller sample sizes and fewer migration-related measures (25). None of the three data sources measures immigrants’ legal status.

Other nationally-representative datasets, such as the Survey of Income and Program Participation (SIPP), the Early Childhood Longitudinal Study program (ECLS), and the National Immunization Surveys (NIS) have been underutilized. These sources have unexploited potential to advance this literature. For example, SIPP measures immigrants’ legal status. ECLS and SIPP provide longitudinal data. Longitudinal datasets are important because repeated cross-sectional datasets such as ACS and NHIS, which recruit new samples each year, can produce biased estimates of policy effects if the composition of the sample changes after a policy is implemented. NIS provides detailed data on vaccination uptake, which is particularly relevant as COVID-19 has raised concerns about how anti-immigrant policies may reduce vaccine uptake among marginalized immigrant populations (26).

To understand how immigrant policies impact different immigrant groups, researchers need detailed information about race/ethnicity; country of birth, citizenship, and legal status; geographic location; and outcomes of interest. However, identifying useful datasets is challenging and time-consuming. Researchers must review documentation spanning years or decades to ensure that sample sizes are adequate, necessary geocodes are available, and key variables are measured consistently over time.

### Purpose of this review

The current study creates a comprehensive resource indexing US datasets that can be used to measure the health effects of state and local policies and other contextual factors. In this systematic review, I identify longitudinal and repeated cross-sectional, nationally-representative, individual-level datasets that include geocodes, race/ethnicity, and at least one health-related outcome. This review describes 54 such datasets, many of which have never been used to study the effects of policies on immigrant health.

To help researchers identify datasets in which key variables are measured consistently over time, I describe measures of race, ethnicity, nativity, country of birth, citizenship, language, and geocodes for each year (2005–2021) of each dataset. I also report language(s) of interview for each survey to aid researchers in identifying which immigrant groups may be excluded due to language barriers. Immigrants who are not proficient in English are often socioeconomically disadvantaged and are more likely to have tenuous legal statuses (27). Thus, surveys conducted only in English will exclude the most vulnerable immigrants (28).

This review is a tool for any researcher who seeks to use large, nationally-representative datasets to study the effects of policy and place on immigrant health. By identifying underutilized datasets that measure health, health behaviors, and health care access, this review should greatly increase the speed and scope of questions that can be answered about the effects of place on immigrant health.

## Methods

This systematic review was conducted and reported following PRISMA guidelines (29). I reviewed all datasets listed at healthdata.gov (30), and datasets related to health compiled by the Department of Health and Human Services (31), the Substance Abuse and Mental Health Services Administration (32), and the National Institute on Alcohol Abuse and Alcoholism (33). To identify relevant datasets indexed by Inter-University Consortium for Political and Social Research (ICPSR), I searched the ICPSR website using the keyword “health.” I also reviewed all datasets included in ICPSR thematic collections related to children and families, health, disability, aging, and demography (34–43). The initial search was run between January 24 – February 9, 2020, and updated December 29 – January 13, 2021. This systematic review was not registered, and no protocol was published.

In reviewing datasets, I applied the following inclusion criteria: nationally-representative sample of the general population (not a specific disease or public benefits sample); released as person-level data; at least 1,000 respondents per year; at least one measure of health, disability, or health care access; geocodes at the state level or below; and data available (with health-related outcomes) for at least two years since 2005. I did include datasets without immigration variables because these datasets are useful to researchers who seek to understand how immigrant policies affect broader populations, including racial/ethnic groups with high proportions of immigrants.

First, I reviewed dataset titles and abstracts/project summaries. If a dataset met the inclusion criteria, or if any of the criteria could not be ascertained based on the title and abstract, the dataset was included in the next stage of screening. For these datasets, I reviewed all available documentation, including project websites, codebooks, public-release datasets, and technical documentation, to determine whether each dataset met the inclusion criteria and to extract information from datasets that met the inclusion criteria. If any information could not be ascertained from documentation, I contacted dataset administrators to inquire directly.

The data extraction sheet in Appendix A was used to extract information from data documentation. The primary variables of interest were race, Hispanic ethnicity, nativity (foreign-born vs. US-born), country of birth, citizenship, legal status, language, health-related measures, and geocodes. I compared measures of race/ethnicity in each dataset to guidelines from the Office of Management and Budget (OMB) and US Department of Health and Human Services (HHS) (Table 1), which set standards for data collection in federal data sources (44, 45). I also extracted technical details about the design and administration of each dataset, including information about which racial/ethnic groups were oversampled in each year.

**Table 1.** Federal standards for collection of information about race, ethnicity, and primary language^a^.

|  | Office of Management and Budget (OMB)<br>1997 minimum standards | Department of Health and Human Services (HHS)<br>2011 standards |
| --- | --- | --- |
| **Ethnicity** | Asked separately from and before race: Hispanic or Latino, not Hispanic or Latino | Asked separately from and before race: Mexican, Mexican-American, Chicano/a; Puerto Rican; Cuban; another Hispanic, Latino/a or Spanish origin; not Hispanic, Latino/a, or Spanish origin |
| **Race** | Allow respondents to choose multiple: White; Black or African American; American Indian or Alaska Native; Asian; Native Hawaiian or other Pacific Islander | Allow respondents to choose multiple: White; Black or African American; American Indian or Alaska Native; Asian Indian; Chinese; Filipino; Japanese; Korean; Vietnamese; other Asian; Native Hawaiian; Guamanian or Chamorro; Samoan; other Pacific Islander |
| **Language** | None | Required for age 5 and older: “How well do you speak English? Very well, well, not well, or not at all.”<br>Recommended for age 5 and older: “Do you speak a language other than English at home?” If yes: “What is this language?” |

HHS = US Department of Health and Human Services; OMB = Office of Management and budget
<sup>a</sup> Office of Management and Budget and Department of Health and Human Services guidelines create requirements for *data collection*; agencies are not required to *release* detailed data to outside researchers.

**Table 2.**
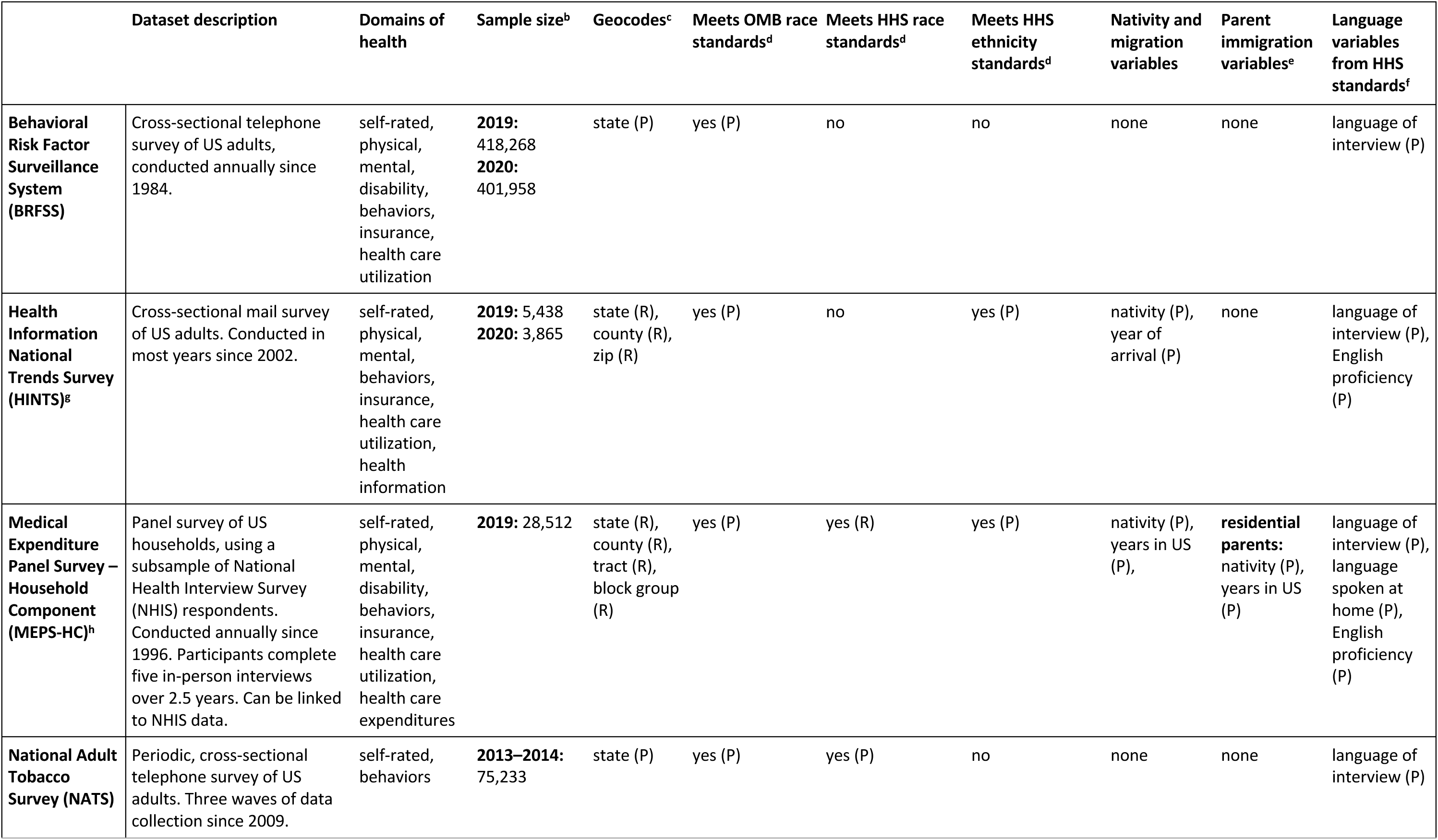
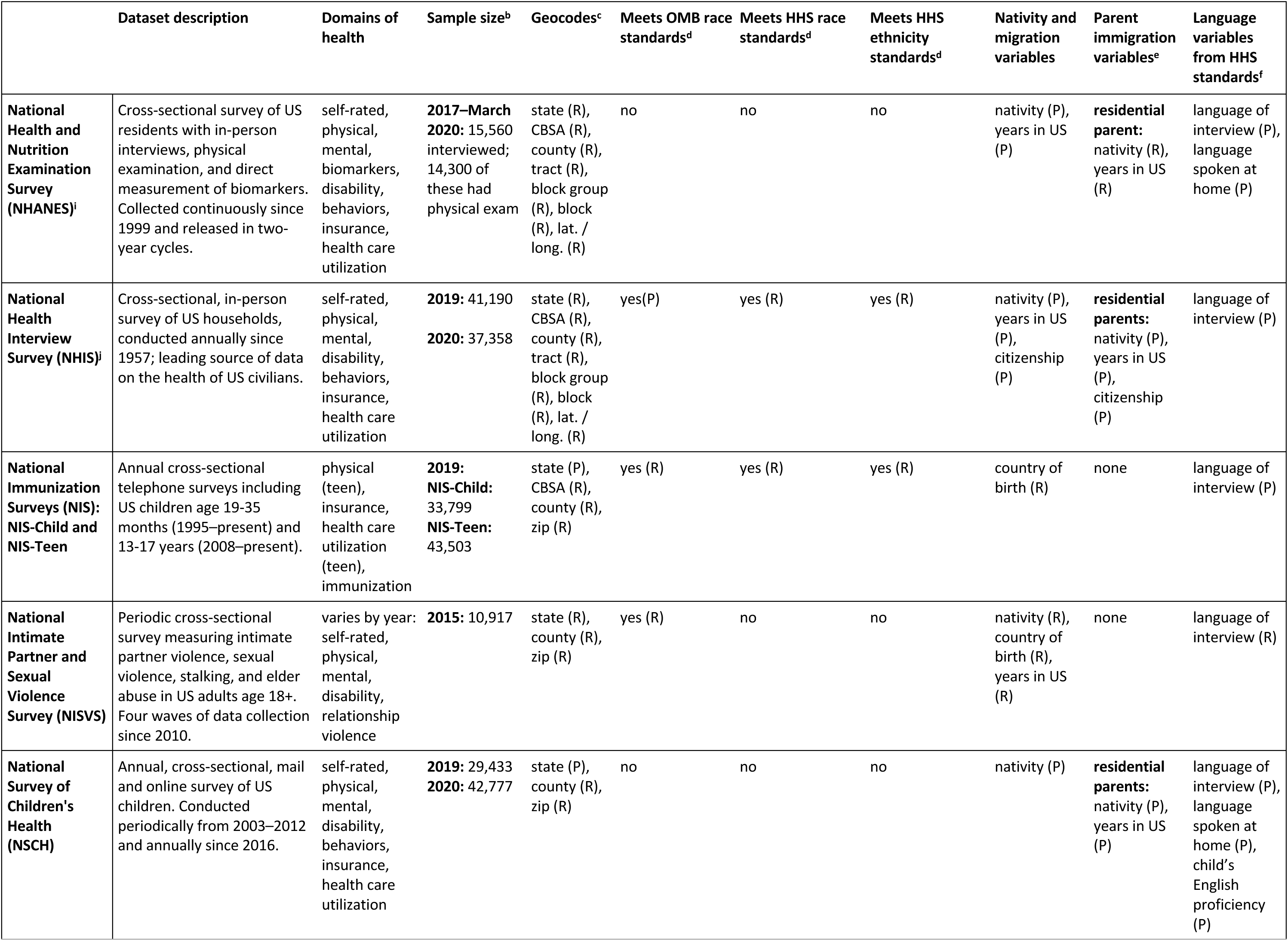
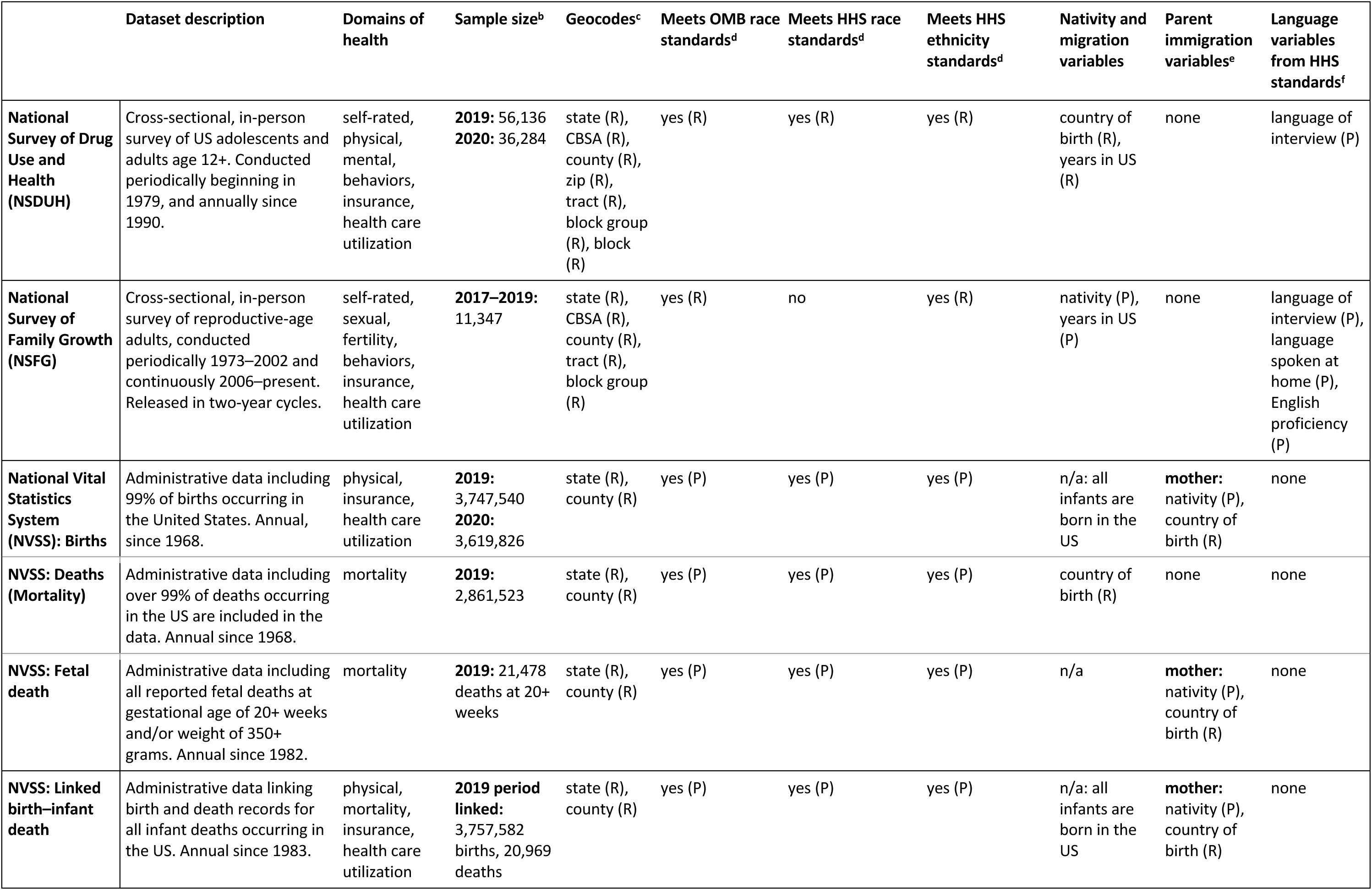
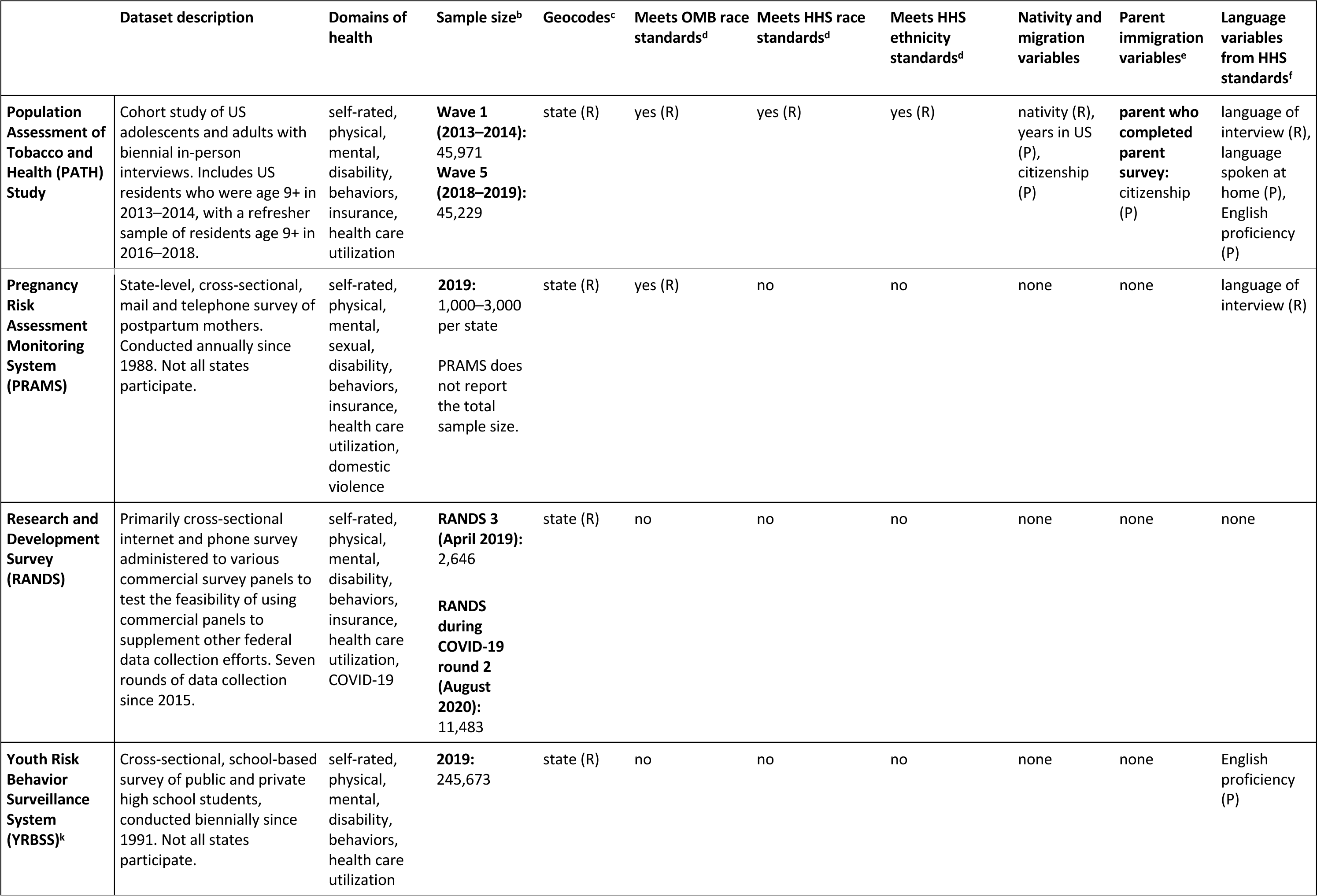
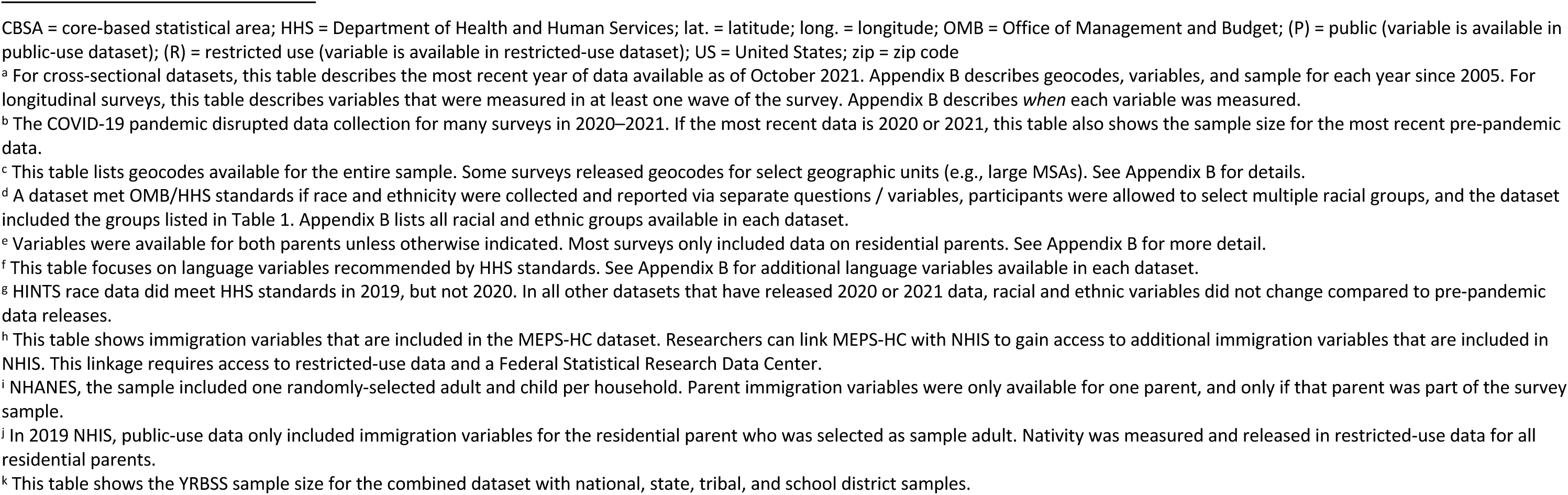
Nationally-representative United States datasets maintained by the US Department of Health and Human Services: Sample sizes and key variables for the most recent year of data for each dataset^a^.

## Results

The searches returned 25,511 records (see flow diagram in Figure 1). After removing duplicates and removing studies that did not meet the criteria, 183 unique datasets were identified for further review. Of these, 54 datasets met the inclusion criteria, including 49 interviews (24, 46–93) and five administrative datasets (89, 94–97). Datasets are described in Tables 3–7 and Appendix B. Five datasets did not include all 50 states in every year, but included most states and were representative of included states (48, 50, 74, 75, 81). The remaining datasets included all 50 states and the District of Columbia. Eight included Puerto Rico (49, 50, 52, 59, 94–97), and six included additional US territories (49, 52, 94–97).

**Figure 1.**
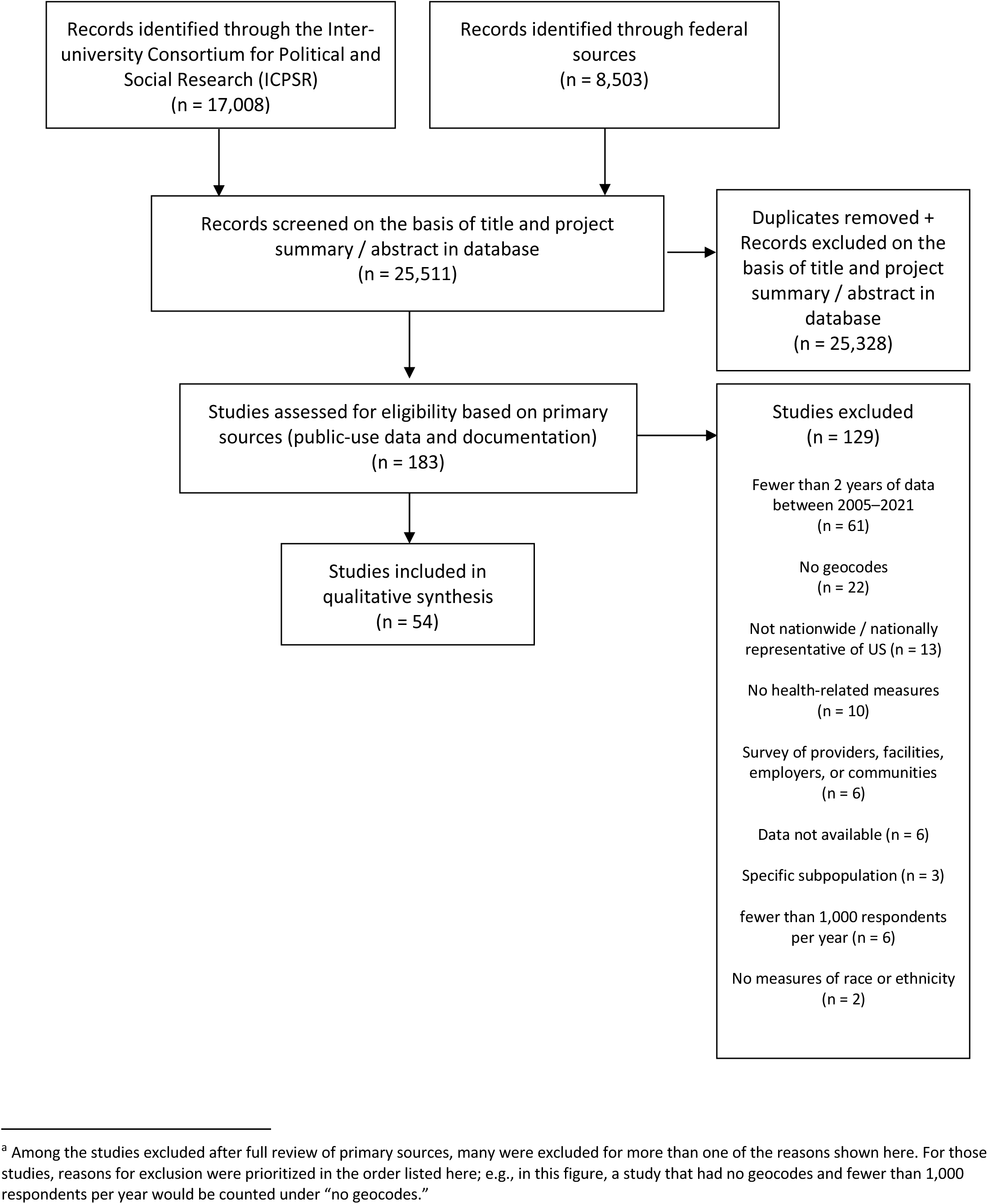
Flow diagram of study search and selection^a^.

**Table 3.** Nationally-representative United States datasets maintained by the US Census Bureau: Sample sizes and key variables for the most recent year of data for each dataset^a^.

|  | Dataset description | Domains of health | Sample size<sup>b</sup> | Geocodes<sup>c</sup> | Meets OMB race standards<sup>d</sup> | Meets HHS race standards<sup>d</sup> | Meets HHS ethnicity standards<sup>d</sup> | Nativity and migration variables | Parent immigration variables<sup>e</sup> | Language variables from HHS standards<sup>f</sup> |
| --- | --- | --- | --- | --- | --- | --- | --- | --- | --- | --- |
| **American Community Survey (ACS)<sup>g</sup>** | Annual, cross-sectional, online survey of US households conducted from 2005–present. | disability, insurance | **2019:** 3,239,553 | state (P), CBSA (P), county (P), zip (R), tract (R), block group (R) | yes (P) | yes (P) | yes (P) | country of birth (P), years in US (P), citizenship (P) | **residential parents of children age <18:** nativity (P)<br>**householder**: country of birth (R), years in US (R), citizenship (R) | language spoken at home (P), English proficiency (P) |
| **American Housing Survey (AHS)** | Panel survey of occupied and vacant residential housing units in the US, 1984–present. Participants complete biennial in-person interviews; duration of follow-up varies. | disability, housing conditions | **2019:** 124,135 | state (R), county (R), tract (R), block group (R), block (R), lat. / long. (R) | yes (P) | yes (P) | no | country of birth (P), years in US (P), citizenship (P) | **residential parents:** country of birth (P), years in US (P), citizenship (P) | language of interview (P) |
| **Arts Basic Survey (ABS)** | The ABS is a supplement to the February Current Population Survey. Sponsored by the National Endowment for the Arts, the ABS collects data on US adults' participation in various artistic activities, performances, and events. ABS has been conducted every one to two years since 2013. | disability | **2018:** 18,116<br>**2020:** 34,995 | state, CBSA, Consolidated Statistical Area, county | yes (P) | no | yes (P) | nativity (P), country of birth (P), citizenship (P), years in US (P) | **all parents:** country of birth<br>**residential parents:** country of birth (P), years in US (P), citizenship (P) | none |
| **Current Population Survey Annual Social and Economic Supplement (CPS-ASEC)** | Annual panel survey of US households, conducted annually since 1962. Participants complete eight in-person or phone interviews over 16 months. This article describes the Annual Social and Economic Supplement (ASEC). | self-rated, disability, insurance | **2019:**<br>180,101<br>**2021:**<br>163,543 | state (P), CBSA (P), county (P), zip (R), tract (R), block group (R), block (R) | yes (P) | yes (P) | yes (P) | country of birth (P), years in US (P), citizenship (P) | **residential parents:**<br>country of birth (P), years in US (P), citizenship (P) | none |
| **Household Pulse Survey (HPS)** | The Household Pulse Survey measures the impacts of the COVID-19 pandemic on the social and economic wellbeing of US households. Data has been collected biweekly for most weeks since the pandemic began. 40 waves of data have been collected so far. Data collection will continue at least through February 2021. | physical, mental, disability, insurance, health care utilization, COVID diagnosis, COVID vaccination, COVID-related behaviors | **Week 39 (September 29 – October 11 2021):**<br>57,064 | state (P) | yes (R) | yes (R) | yes (R) | none | none | none |
| **National Longitudinal Mortality Study (NLMS)** | Research database that links death record data (through 2013) with cross-sectional data from 39 cohorts of from CPS respondents and the 1980 Census. No new cohorts will be added, but the Census Bureau will continue to update mortality data for existing cohorts. | self-rated, disability, mortality, insurance | **March 2011 CPS-ASEC cohort:**<br>130,061 | state (P), county (R), tract (R) | yes (R) | no | yes (R) | region of birth (P), country of birth (R), citizenship (R) | none | none |
| **Survey of Income and Program Participation (SIPP)** | Panel survey of US civilians age 15+. Conducted since 1984. Currently, participants complete annual in-person interviews for four years. | self-rated, fertility, disability, insurance, health care utilization, health care expenditures | **2018 panel, Wave 1:** 763,186<br>**2019 panel, Wave 1 (discontinued after Wave 1):** 593,604 | state (P), CBSA (R), county (R), census place (R), tract (R) | yes (P) | yes (R) | yes (R) | region of birth (P), country of birth (R), years in US (P), citizenship (P), legal status at entry (P), current legal status (R), year acquired LPR status (R) | region of birth (P), **residential parents:** country of birth (R), year of arrival (P), citizenship (P), legal status at entry (P), current legal status (R), year acquired LPR status (R) | language of interview (R), language spoken at home (P), English proficiency (P) |

CBSA = core-based statistical area; HHS = Department of Health and Human Services; lat. = latitude; long. = longitude; OMB = Office of Management and Budget; (P) = public (variable is available in public-use dataset); (R) = restricted use (variable is available in restricted-use dataset); US = United States; zip = zip code
<sup>a</sup> For cross-sectional datasets, this table describes the most recent year of data available as of October 2021. Appendix B describes geocodes, variables, and sample for each year since 2005. For longitudinal surveys, this table describes variables that were measured in at least one wave of the survey. Appendix B describes *when* each variable was measured.
<sup>b</sup> The COVID-19 pandemic disrupted data collection for many surveys in 2020–2021. If the most recent data is 2020 or 2021, this table also shows the sample size for the most recent pre-pandemic data.
<sup>c</sup> This table lists geocodes available for the entire sample. Some surveys released geocodes for select geographic units (e.g., large MSAs). See Appendix B for details.
<sup>d</sup> A dataset met OMB/HHS standards if race and ethnicity were collected and reported via separate questions / variables, participants were allowed to select multiple racial groups, and the dataset included the groups listed in Table 1. Appendix B lists all racial and ethnic groups available in each dataset.
<sup>e</sup> Variables were available for both parents unless otherwise indicated. Most surveys only included data on residential parents. See Appendix B for more detail.
<sup>f</sup> This table focuses on language variables recommended by HHS standards. See Appendix B for additional language variables available in each dataset.
<sup>g</sup> ACS indicated family relationships between the householder and each household member, but not between other household members. Parent immigration variables were available for one parent if that parent was identified as the householder.

**Table 4.** Nationally-representative United States datasets maintained by the Bureau of Labor Statistics: Sample sizes and key variables for the most recent year of data for each dataset^a^.

|  | Dataset description | Domains of health | Sample size<sup>b</sup> | Geocodes<sup>c</sup> | Meets OMB race standards<sup>d</sup> | Meets HHS race standards<sup>d</sup> | Meets HHS ethnicity standards<sup>d</sup> | Nativity and migration variables | Parent immigration variables<sup>e</sup> | Language variables from HHS standards<sup>f</sup> |
| --- | --- | --- | --- | --- | --- | --- | --- | --- | --- | --- |
| **American Time Use Survey (ATUS)<sup>g</sup>** | Annual, cross-sectional, telephone survey of US residents age 15+. Began in 2003 and can be linked to Current Population Survey. | varies by year: self-rated, physical, mental, behaviors | **2019:** 9,435<br>**2020:** 8,782 | state (P) | yes (P) | no | yes (P) | none | none | language of interview (P) |
| **Consumer Expenditure Surveys (CE)** | Annual surveys of US households. Participants complete quarterly in-person or phone interviews for one year. Conducted periodically beginning in 1960 and annually since 1996. Information presented here is for the Interview Survey. | insurance, health care expenditures | **2019:** 11,598<br>**2020:** 20,245 | state (P), CBSA (P), county (R), zip (R), tract (R), block group (R), block (R) | yes (P) | yes (R) | yes (P) | none | none | language of interview (P) |
| **National Longitudinal Survey of Youth 1979 (NLSY79)** | Cohort study of US youth born between 1957-1964, who were living in the US in 1979. Data collection occurred annually from 1979–1994 and biennially since 1996. | self-rated, physical, mental, sexual, fertility, disability, behaviors, insurance, health care utilization | **Wave 1 (1979):** 12,686<br>**Wave 28 (2018/2019):** 6,878 | state (R), CBSA (R), county (R), zip (R), tract (R) | yes (P) | no | yes (P) | nativity (P), country of birth (R), year of arrival (P), citizenship (P) | nativity (P), country of birth (R) | language of interview (P) |
| **National Longitudinal Survey of Youth 1979 Child / Young Adult Survey (NLSCYA)** | Cohort study of children born to female NLSY79 respondents, beginning in 1986. In-person or telephone interviews biennially through age 30, every four years after age 30. | self-rated, physical, mental, sexual, fertility, disability, behaviors, insurance, health care utilization | **ever interviewed:** 11,545<br>**2018:** 4,962 | state (R), CBSA (R), county (R), zip (R), tract (R) | yes (P) | no | no | none | **mother, measured in 1990:** nativity (P), country of birth (R), immigration status (P) | language of interview (P), language child usually speaks (P) |
| **National Longitudinal Survey of Youth 1997 (NLSY97)** | Cohort study of US youth born between 1980-1984, who were living in the US in 1997. In-person or telephone interviews (annually 1997–2012, biennially since 2013). | self-rated, physical, mental, sexual, fertility, disability, behaviors, insurance, health care utilization | **Wave 1 (1997):** 8,984<br>**Wave 18 (2017/2018):** 6,734 | state (R), CBSA (R), county (R), zip (R), tract (R) | yes (P) | no | no | nativity (P), country of birth (R), age at arrival (P), citizenship (P), legal status (P) | region of birth (P), years in US (P) | none |

CBSA = core-based statistical area; HHS = Department of Health and Human Services; lat. = latitude; long. = longitude; OMB = Office of Management and Budget; (P) = public (variable is available in public-use dataset); (R) = restricted use (variable is available in restricted-use dataset); US = United States; zip = zip code
<sup>a</sup> For cross-sectional datasets, this table describes the most recent year of data available as of October 2021. Appendix B describes geocodes, variables, and sample for each year since 2005. For longitudinal surveys, this table describes variables that were measured in at least one wave of the survey. Appendix B describes *when* each variable was measured.
<sup>b</sup> The COVID-19 pandemic disrupted data collection for many surveys in 2020–2021. If the most recent data is 2020 or 2021, this table also shows the sample size for the most recent pre-pandemic data.
<sup>c</sup> This table lists geocodes available for the entire sample. Some surveys released geocodes for select geographic units (e.g., large MSAs). See Appendix B for details.
<sup>d</sup> A dataset met OMB/HHS standards if race and ethnicity were collected and reported via separate questions / variables, participants were allowed to select multiple racial groups, and the dataset included the groups listed in Table 1. Appendix B lists all racial and ethnic groups available in each dataset.
<sup>e</sup> Variables were available for both parents unless otherwise indicated. Most surveys only included data on residential parents. See Appendix B for more detail.
<sup>f</sup> This table focuses on language variables recommended by HHS standards. See Appendix B for additional language variables available in each dataset.
<sup>g</sup> Researchers can link ATUS to the same respondents in the CPS data, allowing access to all immigration variables measured in CPS.

**Table 5.**
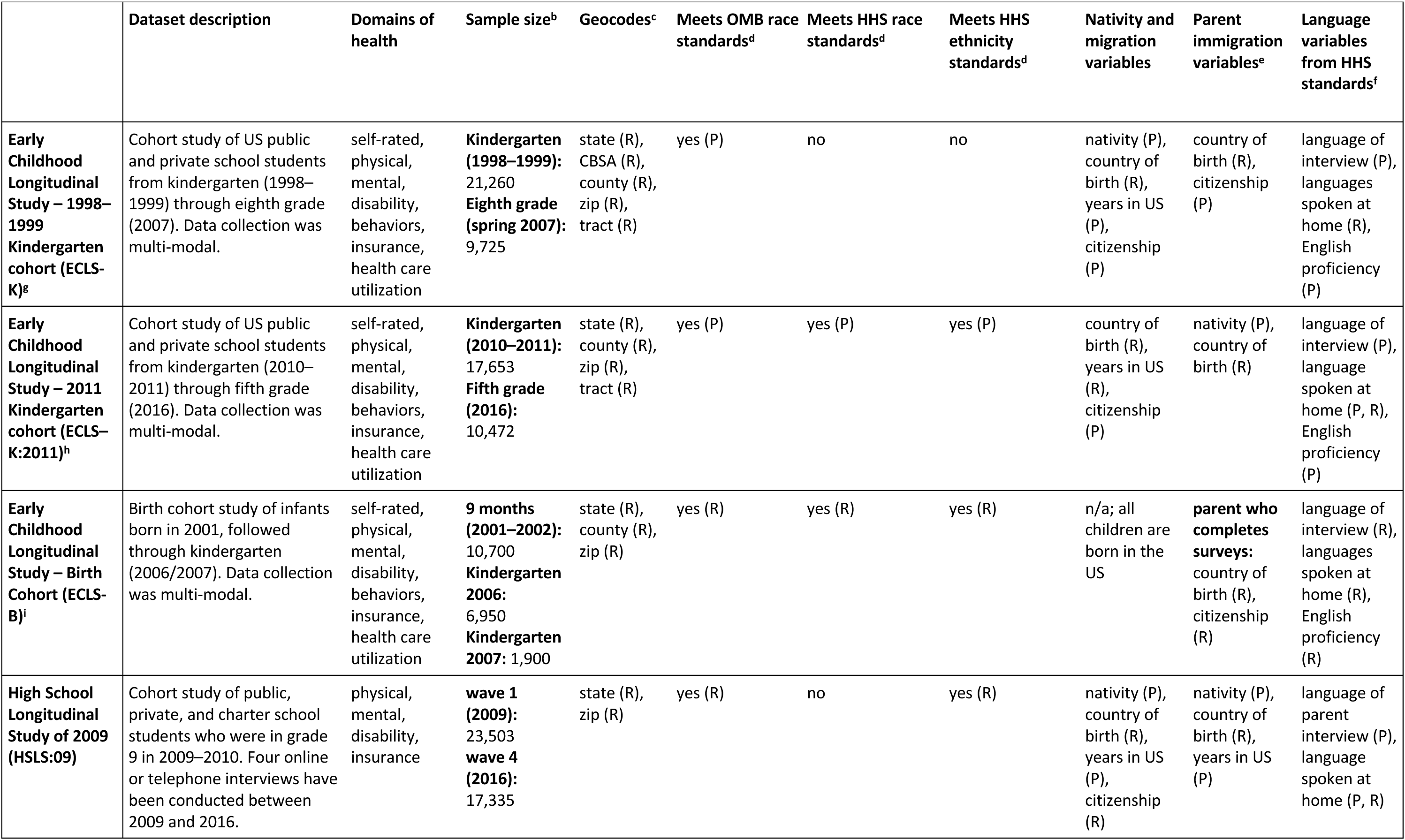
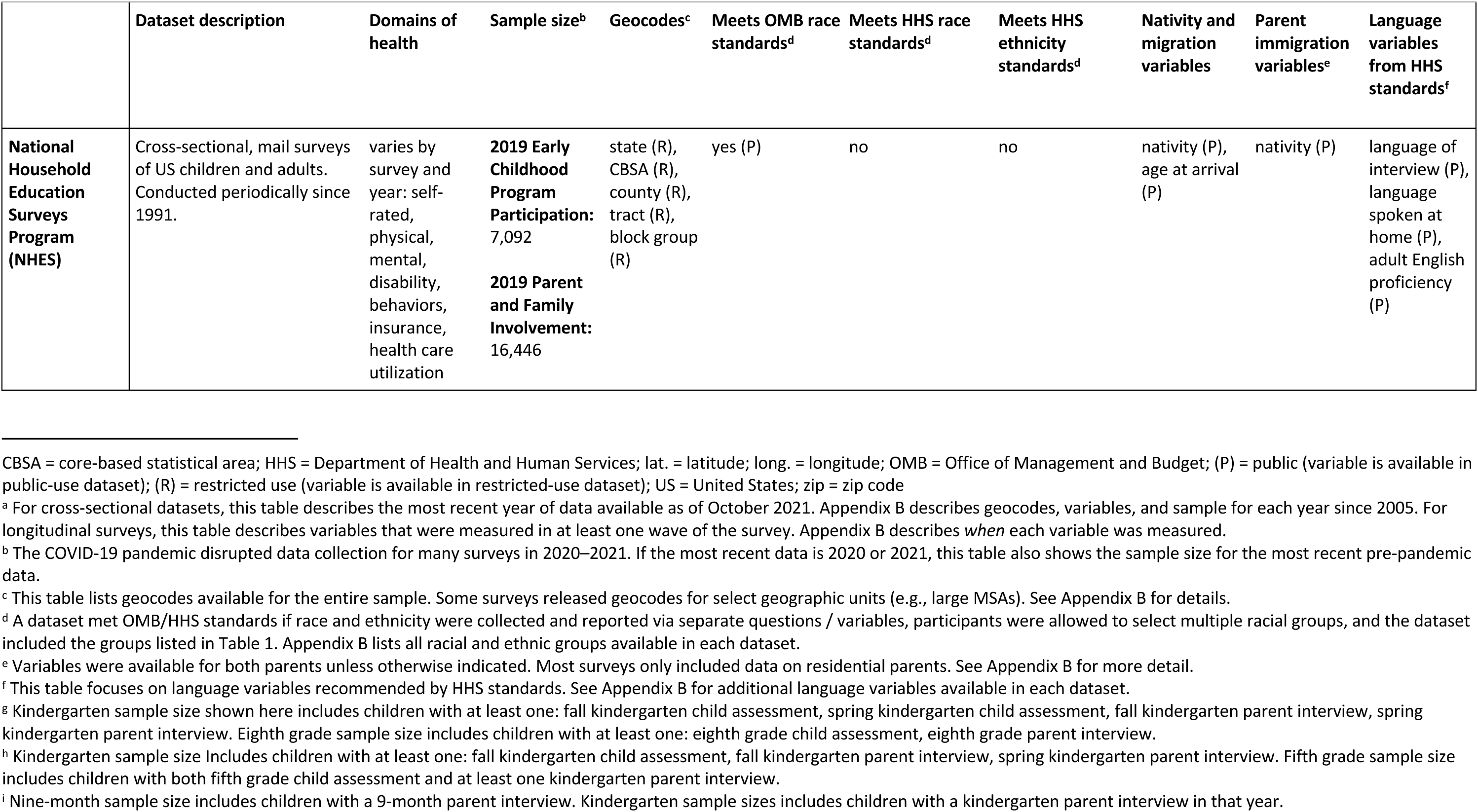
Nationally-representative United States datasets maintained by the US Department of Education: Sample sizes and key variables for the most recent year of data for each dataset^a^.

**Table 6.**
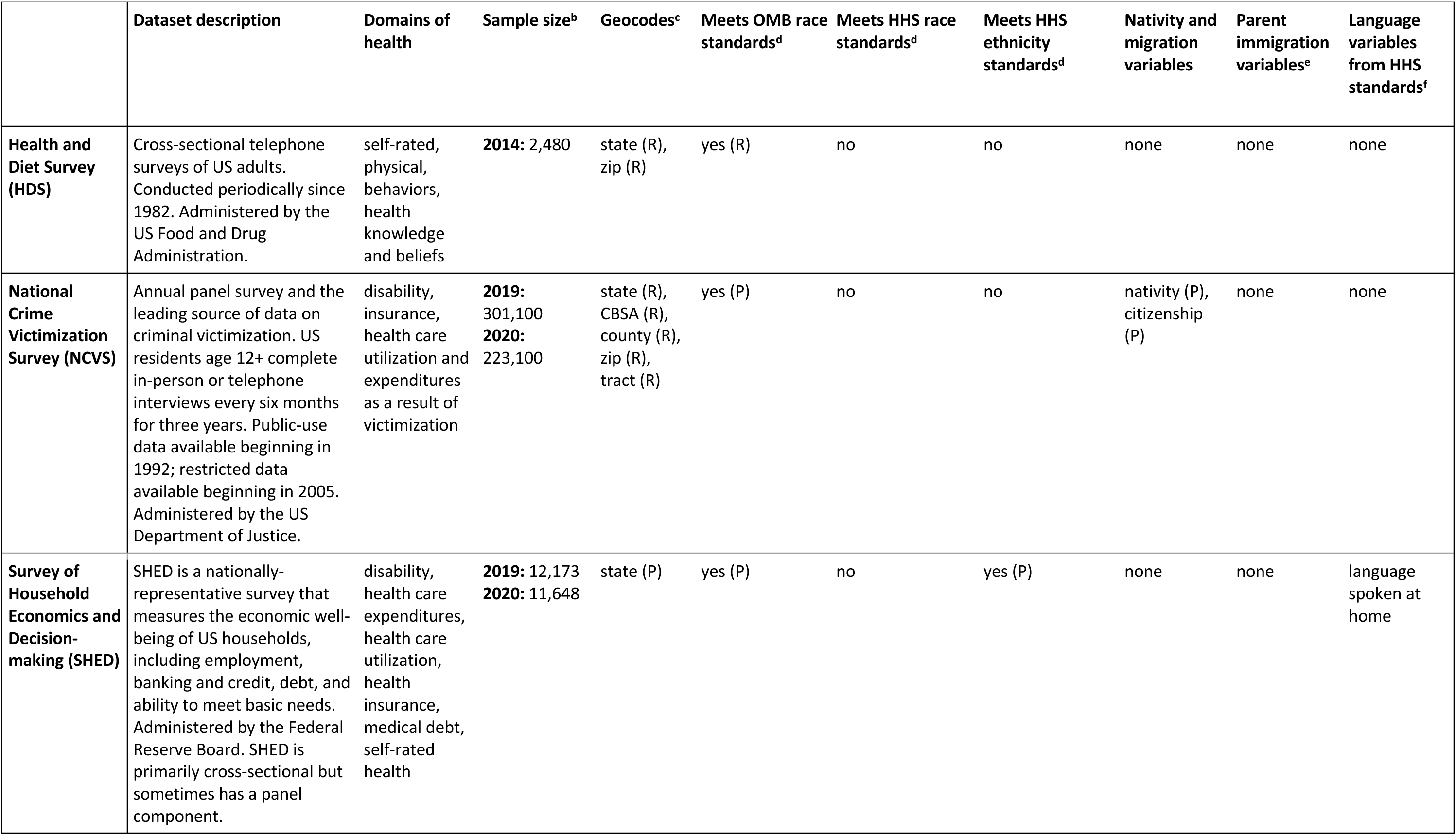
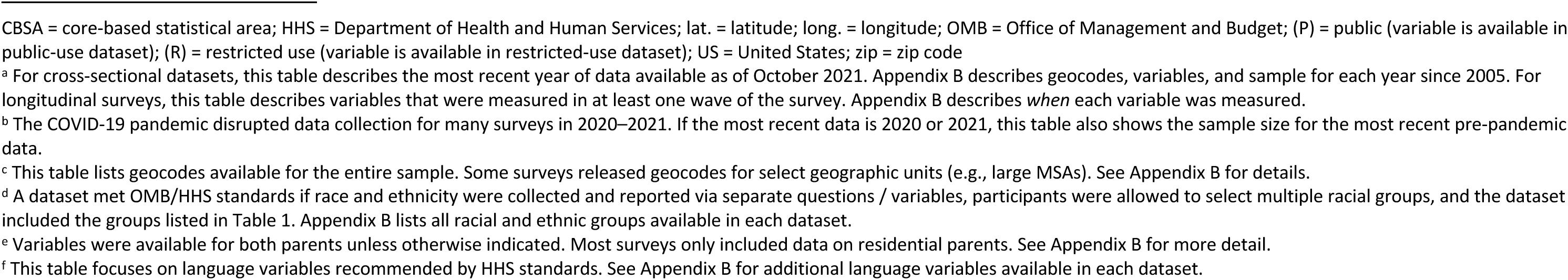
Nationally-representative United States datasets maintained by other federal agencies: Sample sizes and key variables for the most recent year of data for each dataset^a^.

|  | Dataset description | Domains of health | Sample size<sup>b</sup> | Geocodes<sup>c</sup> | Meets OMB race standards<sup>d</sup> | Meets HHS race standards<sup>d</sup> | Meets HHS ethnicity standards<sup>d</sup> | Nativity and migration variables | Parent immigration variables<sup>e</sup> | Language variables from HHS standards<sup>f</sup> |
| --- | --- | --- | --- | --- | --- | --- | --- | --- | --- | --- |
| **Health and Diet Survey (HDS)** | Cross-sectional telephone surveys of US adults. Conducted periodically since 1982. Administered by the US Food and Drug Administration. | self-rated, physical, behaviors, health knowledge and beliefs | **2014:** 2,480 | state (R), zip (R) | yes (R) | no | no | none | none | none |
| **National Crime Victimization Survey (NCVS)** | Annual panel survey and the leading source of data on criminal victimization. US residents age 12+ complete in-person or telephone interviews every six months for three years. Public-use data available beginning in 1992; restricted data available beginning in 2005. Administered by the US Department of Justice. | disability, insurance, health care utilization and expenditures as a result of victimization | **2019:** 301,100<br>**2020:** 223,100 | state (R), CBSA (R), county (R), zip (R), tract (R) | yes (P) | no | no | nativity (P), citizenship (P) | none | none |
| **Survey of Household Economics and Decision-making (SHED)** | SHED is a nationally-representative survey that measures the economic well-being of US households, including employment, banking and credit, debt, and ability to meet basic needs. Administered by the Federal Reserve Board. SHED is primarily cross-sectional but sometimes has a panel component. | disability, health care expenditures, health care utilization, health insurance, medical debt, self-rated health | **2019:** 12,173<br>**2020:** 11,648 | state (P) | yes (P) | no | yes (P) | none | none | language spoken at home |

CBSA = core-based statistical area; HHS = Department of Health and Human Services; lat. = latitude; long. = longitude; OMB = Office of Management and Budget; (P) = public (variable is available in public-use dataset); (R) = restricted use (variable is available in restricted-use dataset); US = United States; zip = zip code
<sup>a</sup> For cross-sectional datasets, this table describes the most recent year of data available as of October 2021. Appendix B describes geocodes, variables, and sample for each year since 2005. For longitudinal surveys, this table describes variables that were measured in at least one wave of the survey. Appendix B describes *when* each variable was measured.
<sup>b</sup> The COVID-19 pandemic disrupted data collection for many surveys in 2020–2021. If the most recent data is 2020 or 2021, this table also shows the sample size for the most recent pre-pandemic data.
<sup>c</sup> This table lists geocodes available for the entire sample. Some surveys released geocodes for select geographic units (e.g., large MSAs). See Appendix B for details.
<sup>d</sup> A dataset met OMB/HHS standards if race and ethnicity were collected and reported via separate questions / variables, participants were allowed to select multiple racial groups, and the dataset included the groups listed in Table 1. Appendix B lists all racial and ethnic groups available in each dataset.
<sup>e</sup> Variables were available for both parents unless otherwise indicated. Most surveys only included data on residential parents. See Appendix B for more detail.
<sup>f</sup> This table focuses on language variables recommended by HHS standards. See Appendix B for additional language variables available in each dataset.

**Table 7.** Nationally-representative United States datasets maintained by universities and other research organizations: Sample sizes and key variables for the most recent year of data for each dataset^a^.

|  | Dataset description | Domains of health | Sample size<sup>b</sup> | Geocodes<sup>c</sup> | Meets OMB race standards<sup>d</sup> | Meets HHS race standards<sup>d</sup> | Meets HHS ethnicity standards<sup>d</sup> | Nativity and migration variables | Parent immigration variables<sup>e</sup> | Language variables from HHS standards<sup>f</sup> |
| --- | --- | --- | --- | --- | --- | --- | --- | --- | --- | --- |
| **American National Election Study (ANES)** | Panel survey of voting-age US citizens age 18+, beginning in 1948. Conducted during presidential election years; participants complete one pre-election survey and one post-election survey in-person or online. Maintained by the University of Michigan and Stanford University. | self-rated, insurance, health care utilization | **2016–2020 panel:** 2,839<br>**New 2020 sample:** 5,441 | state (R), county (R), zip (R), tract (R) | yes (R) | no | yes (P) | nativity (P), country of birth (R), years in US (R), year naturalized (R) | number of parents born outside US (P), number of grandparents born outside US (P) | language of interview (P) |
| **Cooperative Election Study (CES)** | Online survey of US adults, conducted annually since 2006. In election years, CCES is a panel survey with one pre-election wave of data collection and one post-election wave. In non-election years, CCES is a cross-sectional survey conducted in the fall. Maintained by Harvard University. | insurance, health care utilization | **2020:** 61,000 | state (P), congressional district (P), county (P), zip (P) | no | no | yes (P) | nativity (P), citizenship (P) | at least one parent born outside US (P), at least one grandparent born outside US (P) | none |
| **Continuity and Change in Contraceptive Use Study (CCCUS)** | Cohort study of US women who were age 18-39 in 2012. Participants completed online surveys every six months from 2012–2014. Maintained by the Guttmacher Institute. | physical, contraceptive use, fertility, insurance, health care utilization | **wave 1 (2012):** 4,634<br>**wave 4 (2014):** 1,842 | state (R) | no | no | no | nativity (R) | none | language of interview (R) |
| **General Social Survey (GSS)** | Biennial, cross-sectional, in-person or telephone survey of English- and Spanish-speaking US adults. Conducted since 1972 and included a panel component from 2006-2014 and 2016–2020. Maintained by the University of Chicago National Opinion Research Center (NORC). | varies by year: self-rated, mental, disability, insurance | **2018:** 2,348<br>**2016–2020 panel:** 1,823<br>**2021:** 4,032 | state (R), county (R), tract (R) | no | no | yes (P) | nativity (P), citizenship (P), plans to naturalize (P) | nativity (P), number of grandparents born outside US (P) | language of interview (P) |
| **Health and Retirement Study (HRS)** | Cohort study of US adults age 50+ with biennial in-person or telephone interviews. Began in 1992 with individuals born between 1931–1941. A new cohort age 51–56 is added every 6 years. Limited to the coterminous US. Maintained by the University of Michigan. | self-rated, physical, mental, disability, cognition, behaviors, insurance, health care utilization, health care expenditures | **2018 (all cohorts):** 17,146 | state (R), county (R), zip (R), tract (R) | yes (R) | no | yes (R) | nativity (P), country of birth (R), years in US (P), citizenship (P), year naturalized (P) | none | language of interview (P), whether language of interview is usually spoken at home (P) |
| **Health Tracking Household Survey (HTHS)** | Cross-sectional telephone survey of US households. Conducted six times between 1996 and 2010. Limited to the coterminous US. Maintained by the Center for Studying Health System Change. | self-rated, physical, mental, disability, insurance, health care utilization | **2010:** 16,671 | state (R), county (R) | no | no | no | years in US (R), citizenship (R) | none | language of interview (R) |
| **Monitoring the Future (MTF)** | Annual school-based survey of 8th, 10th, and 12th graders, beginning in 1976. Primarily cross-sectional, but about 2,450 respondents from each 12th grade class are followed into adulthood. Participating states vary by year. Maintained by the University of Michigan. | physical, mental, disability, behaviors, health care utilization | **2019:** 42,531<br>**2020:** 11,821 | state (R), county (R), zip (R) | no | no | no | none | none | none |
| **National Alcohol Surveys (NAS)** | Cross-sectional telephone survey of US adults, conducted approximately every 5 years from 1964 to present. Data from 2014 and later are only available on consultation with NAS staff. Maintained by the Alcohol Research Group. | self-rated, physical, mental, behaviors, insurance, health care utilization | **2014–2015:** 6,625 | state (P), MSA (R), zip (R) | yes (P) | no | yes (P) | nativity (P), country of birth (R), years in US (P) | none | language of interview (P) |
| **National Asian American Survey (NAAS)** | Cross-sectional telephone survey of Asian / Asian American adults in the US. Conducted once in 2008 and twice in 2016 (once pre-election and once post-election). Collected by the University of California, Riverside and archived at ICPSR. | self-rated | **2016 pre-election:** 4,787<br>**2016 post-election:** 6,448 | state (R) | no | no | no | country of birth (P), years in US (P), age at arrival (P), citizenship (P), legal status (P) | nativity (P) | language of interview (P), English proficiency (P) |
| **National Poll on Healthy Aging (NPHA)** | Cross-sectional survey of US adults age 50-80, conducted 2-3 times per year since 2017. Maintained by the University of Michigan. | varies by year: self-rated, physical, behaviors, insurance, health care utilization | **May 2019:** 2,966 | state (P) | no | no | no | none | none | none |
| **National Survey of Early Care and Education (NSECE)** | Cross-sectional dataset measuring the availability of, need for, and use of early childhood care and education. Includes a household survey (described in this article) as well as surveys of child care providers. Maintained by the University of Chicago National Opinion Research Center (NORC). | insurance | **2019:** 8,576 households with 15,981 children | state (R)<br>county (R) | yes (R) | no | no | nativity (P) | nativity (P),<br>country of birth (R),<br>years in US (R) | language of interview (P),<br>primary language (P) |
| **National Survey of Teen Relationships and Intimate Violence (STRiV)** | Cohort survey of US adolescents who were age 10-18 in 2013. Participants have completed six annual online surveys since 2013. Only Waves 1–4 are currently available to researchers. Maintained by the University of Chicago National Opinion Research Center (NORC). | mental, behaviors, domestic violence | **wave 1 (2013):** 2,645<br>**wave 4 (2017):** 1,646 | state (R) | no | no | yes (R) | none | none | language of interview (R),<br>primary language (R) |
| **Panel Study of Income Dynamics (PSID)<sup>g</sup>** | Panel study of US families with biennial telephone interviews. Began in 1968 with a sample of 4,802 families. Refresher samples of recent immigrants were added in 1997 (511 families) and 2017 (1,624 families). Maintained by the University of Michigan. | self-rated, physical, mental, disability, insurance, health care utilization, health care expenditures | **Wave 1 (1968):** 4,802 households<br>**Wave 41 (2019):** 9,569 households | state (P),<br>CBSA (R),<br>county (R),<br>zip (R),<br>tract (R),<br>block group (R), block (R) | yes (P) | yes (R) | yes (P) | **reference person and spouse:**<br>nativity (P),<br>country of birth (R) | nativity (P);<br>world region where parent now lives (P)<br>**parent in survey as reference person or spouse:**<br>nativity (P),<br>country of birth (R) | language of interview (P) |
| **Survey of American Family Finances (SAFF)** | Conducted and maintained by Pew Charitable Trusts. | insurance, health care utilization, medical debt, paid sick days | **Wave 1 (2014):** 7,845<br>**Wave 2 (2015):** 5,661 | state (P) | yes (P) | yes (P) | yes (P) | nativity (P), citizenship (P) | none | language of interview (P) |
| **Stress in America survey** | Annual, cross-sectional, online survey of US residents conducted since 2007. Has had a different focus each year, and target population (e.g., defined by age or race) varies by year. Maintained by the American Psychological Association. | self-rated, physical, mental, stress, disability, behaviors, insurance, health care utilization | **2018:** 4,550 | state (R), MSA (R), zip (R) | yes (R) | no | yes (R) | nativity (R), years in US (R), citizenship (R), legal status (R) | number of parents born outside US (R) | language of interview (R), language usually speak at home (R), English proficiency (R) |

CBSA = core-based statistical area; HHS = Department of Health and Human Services; ICPSR = Inter-university Consortium for Political and Social Research; lat. = latitude; long. = longitude; OMB = Office of Management and Budget; (P) = public (variable is available in public-use dataset); (R) = restricted use (variable is available in restricted-use dataset); US = United States; zip = zip code
<sup>a</sup> For cross-sectional datasets, this table describes the most recent year of data available as of October 2021. Appendix B describes geocodes, variables, and sample for each year since 2005. For longitudinal surveys, this table describes variables that were measured in at least one wave of the survey. Appendix B describes *when* each variable was measured.
<sup>b</sup> The COVID-19 pandemic disrupted data collection for many surveys in 2020–2021. If the most recent data is 2020 or 2021, this table also shows the sample size for the most recent pre-pandemic data. 2020 sample sizes are shown for ANES and CES because these are election surveys with the largest samples in presidential election years.
<sup>c</sup> This table lists geocodes available for the entire sample. Some surveys released geocodes for select geographic units (e.g., large MSAs). See Appendix B for details.
<sup>d</sup> A dataset met OMB/HHS standards if race and ethnicity were collected and reported via separate questions / variables, participants were allowed to select multiple racial groups, and the dataset included the groups listed in Table 1. Appendix B lists all racial and ethnic groups available in each dataset.
<sup>e</sup> Variables were available for both parents unless otherwise indicated. Most surveys only included data on residential parents. See Appendix B for more detail.
<sup>f</sup> This table focuses on language variables recommended by HHS standards. See Appendix B for additional language variables available in each dataset.
<sup>g</sup> PSID is a multi-generational study. Some children continue to be followed after they move out of their original household. If those children are head of their new household, nativity and country of birth are recorded for those individuals. Information about their parents can be accessed (if their parents were household head or spouse) by linking family members.

Twenty-seven datasets were designed specifically to collect health data, including vital statistics, health, health care, and/or health behaviors (25, 46–58, 74, 75, 81–84, 89, 93–98). The remaining datasets focused on demographics (59), life course trajectories (66–68), housing (61), economics and labor force (60, 62, 64, 85, 90, 91), education (65, 69–72, 92), violence exposure (73, 78, 88), time use (63), public opinion (76, 77, 79, 80), the arts (87), or experiences during the COVID-19 pandemic (86).

Tables 3–7 show the types of health-related outcomes measured in each dataset. Health insurance coverage was the most common health-related measure (25, 46, 47, 49, 50, 52–57, 59, 60, 62, 64–73, 75–77, 79, 81–86, 89–93, 98). Other common measures included: self-rated or parent-rated health (25, 46–51, 53–58, 60, 62, 63, 65–71, 75–77, 80, 81, 83–85, 88–90, 93, 98); physical health (25, 47–50, 52–58, 63, 65–72, 74, 75, 81–86, 88, 89, 93–98); mental health (25, 47–50, 53–57, 63, 65–72, 74, 75, 77, 78, 81, 83–86, 88, 93); disability (25, 47–50, 53, 54, 57, 59–62, 65–75, 77, 81, 84–90, 92, 93); health behaviors (25, 46–51, 53–58, 63, 65–71, 74, 75, 78, 83, 84, 86, 93, 98); and health care access/utilization (25, 46–50, 52–57, 62, 65–71, 73–76, 79, 81–86, 90, 91, 93–96, 98).

Twenty-seven datasets were repeated cross-sectional (25, 46–52, 54–56, 58, 59, 63, 65, 80, 81, 83, 84, 86–88, 92, 94, 96–98). Twenty-two were longitudinal (53, 57, 60–62, 64, 66–73, 75, 76, 78, 82, 85, 89, 91, 95), with follow-up ranging from a few weeks (76) to over 50 years (85). Five datasets included both longitudinal and cross-sectional components (74, 77, 79, 90, 93). For 24 datasets, data were released annually (25, 49, 50, 52–55, 59, 62–64, 73, 74, 84, 90, 91, 94–97) or more frequently (60, 82, 86, 98). The remaining surveys released data less frequently. Nine surveys have concluded data collection (69–71, 78, 80–82, 89, 91).

Sample sizes in the most recent year of each dataset ranged from 1,646 (78) to 3,747,540 (94). In the most recent year of data collection, fourteen surveys oversampled Black respondents (46–48, 56–58, 63, 65–68, 75, 83, 91), sixteen oversampled Hispanic respondents (46–48, 56, 58, 60, 63, 65–68, 75, 83, 87, 89, 91), and six oversampled Asian respondents (47, 69–72, 80). Most datasets had relatively large samples of White, Black, and Hispanic respondents, but small samples of Asian, Pacific Islander, American Indian, and Alaska Native respondents. Most datasets also had relatively small samples of non-Mexican Hispanic respondents. The exceptions were birth records, death records, and the American Community Survey (ACS), which had very large sample sizes (2,500,000–4,000,000 respondents in the most recent year).

### Race and ethnicity

Tables 3–7 give information about race, ethnicity, nativity, migration, language, and geocodes for the most recent year of each survey. Appendix B provides detailed information about each survey for each year from 2005–2021. In the most recent year, 42 datasets met or exceeded OMB standards for race and ethnicity (25, 46, 49–53, 55–73, 75, 76, 83–92, 94–97). Twenty met or exceeded HHS standards for race (25, 51–53, 55, 57, 59–62, 64, 69, 70, 85, 86, 91, 94–97), and 33 met or exceeded HHS standards for ethnicity (25, 46, 52, 53, 55–57, 59, 60, 62–64, 66, 69, 71, 72, 75–79, 83–87, 89–91, 94–97).

Smaller datasets released limited detail about race and ethnicity to protect participant confidentiality. In many cases, broad racial and ethnic groups were available in public-use data, while more detailed categories were available in restricted-use data. In general, measures of race and ethnicity have become more detailed over time.

### Migration

Two-thirds of datasets included immigration variables. Thirty-five included nativity (25, 46, 47, 52–57, 59–62, 65, 66, 68, 70–73, 75–77, 79, 80, 82–85, 87–89, 91, 92, 97). Twenty-one indicated country of birth (52, 55, 59–62, 66, 68, 70–72, 75, 76, 80, 83, 85, 87–89, 92, 97). Twenty-five provided information about how long the respondent had lived in the US (25, 46, 47, 53, 55–57, 59–62, 65, 66, 68, 70–72, 75, 76, 80, 81, 83, 84, 87, 88). Twenty-three identified whether the respondent was a US citizen (25, 57, 59–62, 66, 68, 70–73, 75–77, 79–81, 84, 85, 87, 89, 91). Only four indicated legal status for noncitizens (62, 68, 80, 84).

Half of studies included migration variable(s) for one or both parents, typically residential parents of minor children. Twenty-seven released parent nativity (25, 47, 53, 54, 59–62, 65–72, 76, 77, 79, 80, 84, 85, 87, 92, 94–96), 16 released country of birth (59–62, 66, 67, 69–72, 85, 87, 92, 94–96), 12 released how long the parent had lived in the US (25, 47, 53, 54, 59–62, 68, 72, 87, 92), nine released citizenship (25, 57, 59–62, 69, 70, 87), and two released legal status (62, 67).

### Language

Of the 49 surveys, four were conducted only in English (48, 74, 79, 93). Twenty-five collected data in English and Spanish (46, 47, 49–51, 55–58, 63, 65, 72, 75–78, 82–84, 86, 88, 90–92, 98). The remaining surveys used one or more of the following methods to collect data in additional languages: translated written interview/survey documents (70, 80), used professional interpreters by phone or in-person (25, 52, 54, 62, 69, 71, 73), utilized bilingual interviewers (25, 59, 60, 62, 64, 66–68, 70, 80, 81, 85, 87), and/or asked family members or neighbors to interpret (25, 53, 59–62, 64, 73, 87). In accordance with HHS guidelines, 15 datasets included information about English proficiency (46, 48, 53–57, 59, 62, 65, 69–71, 80, 84), and 17 indicated the primary language(s) that respondents spoke at home (46, 47, 53, 54, 57, 59, 62, 65,69–72, 75, 78, 84, 90, 92).

### Geocodes

All datasets identified respondents’ state of residence. Eighteen identified core-based statistical area (CBSA), metropolitan statistical area (MSA), or city (25, 47, 52, 55, 59, 60, 62, 64–68, 70, 73, 83–85, 87). Thirty-seven identified county (25, 46, 47, 52–56, 59–62, 64–71, 73–77, 79, 81, 85–89, 92, 94–97), and twenty identified zip code (52, 54–56, 58–60, 64, 69–75, 79, 83–85, 88). Over one-third of datasets identified sub-county geocode(s), including census tract (25, 46, 47, 53, 55, 59–62, 64, 65, 70, 71, 73, 75–77 ,85, 89), block group (25, 46, 47, 53, 55, 59, 64, 65, 85), block (25, 47, 55, 59, 64, 85), census place (25, 47, 59–62), and latitude / longitude (25, 47).

In most datasets, sub-state geocodes, country of birth, and legal status were available only in restricted-use datasets. Appendix B describes data access and associated fees for restricted-use data.

### Impacts of COVID-19 on data collection

In April 2020, the US Census Bureau began the Household Pulse Survey to monitor the social and economic effects of the COVID-19 pandemic. Data about health, education, economic well-being, and COVID-19 diagnoses and vaccines are available for most weeks since April 2020 (86).

The COVID-19 pandemic disrupted data collection for many other surveys. Early in the pandemic, workplaces closed and employees worked remotely from home. Call centers closed, delaying data collection for telephone surveys. The switch to remote work also delayed mailing of surveys and other materials to participants (99).

Surveys conducted through in-person interviews were most impacted. Some quickly pivoted to mail, internet, or telephone; others suspended operations until it was safe to return to in-person data collection. The switch to remote data collection led to lower response rates and prevented collection of biospecimens (99). Some surveys have returned to in-person data collection (57, 59); others remained at least partially remote during 2021 (25, 77).

The pandemic led to higher-than-usual levels of nonresponse bias in the American Community Survey (ACS) and the Current Population Survey (CPS). The Census Bureau cautions researchers that in 2020 ACS data, weighting cannot adequately adjust for nonresponse bias (100). Nonresponse bias also affects 2020 CPS (101) and Consumer Expenditure Surveys (102) data, to a lesser extent than ACS. In 2020, nonresponse bias led to underrepresentation of low-socioeconomic status, Hispanic, foreign-born, and noncitizen individuals (101). Researchers should expect similar nonresponse bias in other 2020 surveys.

## Discussion

State and local immigrant policies are associated with self-rated health and health care access for Hispanic families (3). Much remains unknown about how these policies affect non-Hispanic immigrants, or to what extent the policies are associated with objective measures of health. To fill these gaps, researchers need nationally-representative data sources that include a broad range of racial/ethnic and immigrant groups and are collected over a period of years or decades. This systematic review identified 54 such datasets.

Overall, major strengths included: (1) data collection about race and ethnicity has improved over time, in response to OMB and HHS guidelines; (2) nativity was available in two-thirds of datasets, while country of birth, citizenship, and parent nativity were each available in about half; (3) most surveys were translated into Spanish, with interviews conducted by Spanish-speaking interviewers; (4) over two-thirds of the datasets included county and sub-county geocodes, which will facilitate research on the effects of county/municipal policies; and (5) almost 75% of datasets included specific measures of physical and/or mental health. Over recent years, many researchers have gained access to restricted federal data due to the expansion of Federal Statistical Research Data Centers (FSRDCs) to thirty-one locations nationwide (103).

The datasets also had some limitations in common. (1) While over one-quarter of data sources included oversamples of Hispanic and Black respondents, few included Asian oversamples, and none oversampled foreign-born respondents. As a result, data sources had relatively small sample-sizes of non-Hispanic immigrants. (2) Only five datasets measured legal status, which limits researchers’ ability to study policy effects on undocumented immigrants. (3) Fewer than half of surveys were administered in languages other than Spanish, leading to underrepresentation of immigrants from countries outside the Americas.

Dataset and survey administrators could make several changes to improve the quality of data available for immigrant populations. First, oversampling of Asian, non-Mexican Hispanic, and/or foreign-born respondents would provide larger sample sizes for understudied immigrant groups. Second, although professional translation and trained interpreters are costly, these resources should be utilized to improve data collection for immigrants who speak neither English nor Spanish (104). Third, surveys should collect data on country of birth, citizenship, and legal status when possible. Measurement of legal status is important in order to improve our understanding of how policy impacts differ by legal status. However, survey administrators should consider both the risks and benefits of collecting data about immigration status (21, 105).

Some of the datasets can be linked to each other or to additional sources to provide additional individual-level data about participants. For example, NHIS can be linked to MEPS, mortality, Housing and Urban Development, and Medicare and Medicaid data (106). NLSY79 and NLSY Children and Young Adults can be linked so that researchers have detailed longitudinal data about mothers and their children (66, 67). The Census Bureau links individuals over time across different years of Census and administrative data, creating a longitudinal dataset (107). These linked data sources will provide researchers with more detailed and longitudinal data.

Data about immigrants in 2020–2021 surveys may be of lower quality than previous years due to challenges related to the 2020 Census and the COVID-19 pandemic. The Trump administration planned to add a citizenship question to the 2020 Census. Researchers estimated that this would lead to an 8% drop in the response rate for households with noncitizen residents (105). Ultimately, the citizenship question was not included in the Census, but the Trump administration planned to use other government data sources to identify noncitizens in the Census. This effort was halted by the Biden administration in January, 2021 (108). The COVID-19 pandemic also introduced major challenges for data collection. Many surveys likely had lower-than-usual response rates for low-income, Hispanic, and immigrant individuals. Nonresponse bias may lead to significant challenges for immigration research in 2020–2021.

This review has several limitations. First, I included all data sources in the largest and most commonly-used data repositories; there may be additional datasets not included in these repositories. Second, data sources that do *not* fit the inclusion criteria (e.g., state-specific databases and data aggregated to the state or county level) may be useful for specific research questions. Third, while this review provides a full description of demographic and migration measures over time, I do not provide an exhaustive review of how *health* is measured. Such information is outside the scope of this paper; many of the datasets have dozens or hundreds of health-related measures each year. Finally, some data sources described here are not available for every year; researchers studying short-term effects of policies need annual data.

ACS NHIS, and birth records remain the largest, longest-running, and most versatile secondary datasets available for researchers to examine the effects of policy and place on health care access, public benefits uptake, and overall health status. However, many other data sources have unique benefits for specific research questions. For example, MEPS-HC includes detailed, longitudinal (2.5-year) data about health care expenditures and can be linked to NHIS (53). ECLS surveys include directly-measured data on child development (69–71), while NHANES includes biomarkers (47). ECLS (69–71) and NLSY (66–68) are longitudinal surveys with medium-to long-term follow-up of the same individuals. Use of these data sources can help researchers uncover the ways that policy and place affect many physical and mental health outcomes for immigrants from all over the world.

## Supporting information

Appendix A Data extraction form

Appendix B Detailed information about datasets

Appendix C Sample sizes by dataset

## Data Availability

All data used in this study are freely available online at study websites. Supplementary files will be attached to this submission.

## Acknowledgements

As the sole author, I was responsible for conceptualizing, designing, and conducting this study, and preparing the text. There are no conflicts of interest. This manuscript has not been published elsewhere and is not under consideration by another journal. This study was funded by the Eunice Kennedy Shriver National Institute of Child Health and Human Development (grant number 2T32HD049302-11) and National Center for Advancing Translational Sciences (grant number UL1TR002373).

## Reference List

1. Radford J, Noe-Bustamante L. Facts on U.S. immigrants, 2017: Statistical portrait of the foreign-born population in the United States. https://www.pewresearch.org/hispanic/2019/06/03/facts-on-u-s-immigrants/. Published 2019. Accessed March 3, 2020.

2. Urban Institute. The Urban Institute children of immigrants data tool. https://children-of-immigrants-explorer.urban.org/pages.cfm. Published 2022. Accessed January 23, 2022.

3. Perreira KM, Pedroza JM. Policies of exclusion: Implications for the health of immigrants and their children. Annu Rev Public Health. 2019;40:147–166. (doi: 10.1146/annurev-publhealth-040218-044115).

4. Hatzenbuehler ML, Prins SJ, Flake M, et al. Immigration policies and mental health morbidity among Latinos: A state-level analysis. Soc Sci Med. 2017;174:169–178. (doi:10.1016/j.socscimed.2016.11.040).

5. Torche F, Sirois C. Restrictive immigration law and birth outcomes of immigrant women. Am J Epidemiol. 2018;188(1):24–33. (doi:10.1093/aje/kwy218).

6. Wang JSH, Kaushal N. *Health and Mental Health Effects of Local Immigration Enforcement*. Cambridge, MA: National Bureau of Economic Research; 2018. (NBER Working Papers, no. 24487). (doi: 10.3386/w24487).

7. Gelatt J, Koball J, Bernstein H, et al. State Immigration Enforcement Policies: How They Impact Low-Income Households. Washington, DC: Urban Institute; 2017. (Urban Institute Research Report).

8. Pedraza FI, Zhu L. Immigration enforcement and the “chilling effect” on Latino Medicaid enrollment. http://healthpolicyscholars.org/sites/healthpolicyscholars.org/files/pedrazazhu_medicaid.pdf. Published 2013. Accessed May 13, 2016.

9. Nichols VC, LeBrón AMW, Pedraza FI. Spillover effects: Immigrant policing and government skepticism in matters of health for Latinos. Public Adm Rev. 2018;78(3):432–443. (doi:10.1111/puar.12916).

10. Amuedo-Dorantes C, Arenas-Arroyo E, Sevilla A. Immigration enforcement and economic resources of children with likely unauthorized parents. J Public Econ. 2018;158:63–78. (doi:10.1016/j.jpubeco.2017.12.004).

11. Venkataramani AS, Shah SJ, O’Brien R, et al. Health consequences of the US Deferred Action for Childhood Arrivals (DACA) immigration programme: A quasi-experimental study. Lancet Public Health. 2017;2(4):e175--e181. (doi:10.1016/S2468-2667(17)30047-6).

12. Potochnick S, May SF, Flores LY. In-state resident tuition policies and the self-rated health of high-school-aged and college-aged Mexican noncitizen immigrants, their families, and the Latina/o community. Harv Educ Rev. 2019;89(1):1–29.

13. Hainmueller J, Lawrence D, Martén L, et al. Protecting unauthorized immigrant mothers improves their children’s mental health. Science. 2017;357(6355):1041–1044. (doi:10.1126/science.aan5893).

14. Wherry LR, Fabi R, Schickedanz A, et al. State and federal coverage for pregnant immigrants: Prenatal care increased, no change detected for infant health. Health Aff (Millwood). 2017;36(4):607–615. (doi:10.1377/hlthaff.2016.1198).

15. Graefe DR, Hasanali SH, DeJong GF, et al. CHIP-ing away at health disparities: Has state-provided health insurance reduced race- and nativity-based differences in health care utilization among US children? Can Public Policy. 2015;41(S2):S70--S79. (doi:10.3138/cpp2014-072).

16. Morey BN. Mechanisms by which anti-immigrant stigma exacerbates racial/ethnic health disparities. Am J Public Health. 2018;108(4):460–463. (doi: 0.2105/AJPH.2017.304266).

17. De Trinidad Young ME, León-Pérez G, Wells CR, et al. Inclusive state immigrant policies and health insurance among Latino, Asian/Pacific Islander, Black, and White noncitizens in the United States. Ethn Health. 2019;24(8):960–972. (doi:10.1080/13557858.2017.1390074)

18. Migration Policy Institute. Regions of birth for immigrants in the United States, 1960-present. https://www.migrationpolicy.org/programs/data-hub/charts/regions-immigrant-birth-1960-present Published August 14, 2013. Accessed January 29, 2021.

19. De Trinidad Young ME, León-Pérez G, Wells CR, et al. More inclusive states, less poverty among immigrants? An examination of poverty, citizenship stratification, and state immigrant policies. Popul Res Policy Rev. 2018;37:205–228. (doi:10.1007/s11113-018-9459-3).

20. American Immigration Council. How the United States immigration system works. https://www.americanimmigrationcouncil.org/research/how-united-states-immigration-system-works. Published September 14, 2021. Accessed January 25, 2022.

21. Young MEDT, Madrigal DS. Documenting legal status: A systematic review of measurement of undocumented status in health research. Public Health Rev. 2017;38(1):26. (doi:10.1186/s40985-017-0073-4).

22. Van Hook J, Bachmeier JD, Coffman DL, et al. Can we spin straw into gold? An evaluation of immigrant legal status imputation approaches. Demography. 2015;52:329–354. (doi:10.1007/s13524-014-0358-x).

23. Davidoff A. Using the National Health Interview Survey for policy research on health insurance. JMS Proceedings, Section on Health Policy Statistics. Alexandria, VA: American Statistical Association; 2007:1576–1580.

24. Blewett LA, Call KT, Turner J, et al. Data resources for conducting health services and policy research. Annu Rev Public Health. 2018;39(1):437–452. (doi:10.1146/annurev-publhealth-040617-013544).

25. National Center for Health Statistics. About the National Health Interview Survey (NHIS). https://www.cdc.gov/nchs/nhis/about_nhis.htm. Published September 17, 2020. Accessed January 23, 2022.

26. Artiga S, Ndugga N, Pham O. Immigrant access to COVID-19 vaccines: Key issues to consider. https://www.kff.org/racial-equity-and-health-policy/issue-brief/immigrant-access-to-covid-19-vaccines-key-issues-to-consider/. Published January 13, 2021. Accessed December 27, 2021.

27. Batalova J, Fix M. A profile of limited English proficient adult immigrants. Peabody J Educ. 2010;85(4):511–534. (doi:10.2307/25759046).

28. Rodríguez-Lainz A, McDonald M, Penman-Aguilar A, et al. Getting data right – and righteous to improve Hispanic or Latino health. J Healthc Sci Humanit. 2016;6(3):60–83.

29. Page MJ, McKenzie JE, Bossuyt PM, et al. The PRISMA 2020 statement: an updated guideline for reporting systematic reviews. BMJ. 2021;372:n71. (doi:10.1136/bmj.n71).

30. US Department of Health and Human Services (HHS). HealthData.gov. https://healthdata.gov. Updated January 13, 2022. Accessed January 13, 2022.

31. Office of the Assistant Secretary for Planning and Evaluation. Directory of health and human services data resources. Published date unknown. Accessed December 29, 2021. https://aspe.hhs.gov/directory-health-and-human-services-data-resources

32. Dean C, Eaton W, Goettsche E, et al. Sources for behavioral health and health services research data analysis. https://www.samhsa.gov/sites/default/files/topics/data_outcomes_quality/data-compendium.pdf. Published July 22, 2016. Accessed February 2, 2020.

33. Alcohol Epidemiologic Data System (AEDS). Alcohol Epidemiologic Data Directory. Arlington, VA: CSR, Incorporated; 2012.

34. Inter-University Consortium for Political and Social Research (ICPSR). DSDR: Data Sharing for Demographic Research. https://www.icpsr.umich.edu/icpsrweb/content/DSDR/index.html. Published 2022. Accessed January 13, 2022.

35. Inter-University Consortium for Political and Social Research (ICPSR). ADDEP: Archive of Data on Disability to Enable Policy and research: Advancing research on disability. https://www.icpsr.umich.edu/icpsrweb/ADDEP/search/studies. Published 2022. Accessed January 13, 2022.

36. Inter-University Consortium for Political and Social Research (ICPSR). Child & Family Data Archive (CFData). https://www.childandfamilydataarchive.org/cfda/pages/cfda/index.html. Published 2022. Accessed January 13, 2022.

37. Inter-university Consortium for Political and Social Research. Child Care & Early Education Research Connections. https://www.researchconnections.org/childcare/search/studies. Published 2019. Accessed January 25, 2020.

38. Inter-university Consortium for Political and Social Research. HMCA: Health & Medical Care Data Archive: Data archive of the Robert Wood Johnson Foundation. https://www.icpsr.umich.edu/web/HMCA/search/studies. Published 2022. Accessed January 13, 2022.

39. Inter-university Consortium for Political and Social Research. NAHDAP: National Addiction & HIV Data Archive Program. https://www.icpsr.umich.edu/icpsrweb/NAHDAP/search/studies. Published 2022. Accessed January 13, 2022.

40. Inter-University Consortium for Political and Social Research (ICPSR). NACDA: National Archive of Computerized Data on Aging. https://www.icpsr.umich.edu/icpsrweb/NACDA/search/studies. Published 2022. Accessed January 13, 2022.

41. Inter-university Consortium for Political and Social Research. ResearchDataGov: Application portal for restricted data for federal statistics. https://www.icpsr.umich.edu/web/appfed/search/studies. Published 2022. Accessed January 13, 2022.

42. Inter-University Consortium for Political and Social Research (ICPSR). United States Census Bureau Data Repository. https://census.icpsr.umich.edu/census/. Published 2022. Accessed January 13, 2022.

43. Inter-University Consortium for Political and Social Research (ICPSR). RCMD: Resource Center for Minority Data. https://www.icpsr.umich.edu/icpsrweb/RCMD/search/studies. Published 2022. Accessed January 13, 2022.

44. Office of Management and Budget (OMB). Revisions to the standards for the classification of federal data on race and ethnicity. Fed Regist. 1997;62(210):59782–58790.

45. Dorsey R, Graham G, Glied S, et al. Implementing health reform: Improved data collection for the monitoring of health disparities. Annu Rev Public Health. 2014;35:123–138. doi:10.1146/annurev-publhealth-032013-182423

46. National Center for Health Statistics. National Survey of Family Growth (NSFG). https://www.cdc.gov/nchs/nsfg/index.htm. Published 2020. Accessed April 20, 2020.

47. National Center for Health Statistics. National Health and Nutrition Examination Survey (NHANES). https://www.cdc.gov/nchs/nhanes/index.htm. Published 2020. Accessed April 20, 2020.

48. Centers for Disease Control and Prevention. Youth Risk Behavior Surveillance System (YRBSS). Adolescent and School Health. https://www.cdc.gov/healthyyouth/data/yrbs/index.htm. Published 2018. Accessed January 24, 2020.

49. Centers for Disease Control and Prevention. About BRFSS. Behavioral Risk Factor Surveillance System. https://www.cdc.gov/brfss/about/index.htm. Published 2014. Accessed January 23, 2019.

50. Centers for Disease Control and Prevention. PRAMS. https://www.cdc.gov/prams/index.htm. Published 2020. Accessed January 24, 2020.

51. Centers for Disease Control and Prevention. National Adult Tobacco Survey (NATS). https://www.cdc.gov/tobacco/data_statistics/surveys/nats/index.htm. Published 2018. Accessed January 24, 2020.

52. Centers for Disease Control and Prevention. About the National Immunization Surveys (NIS). https://www.cdc.gov/vaccines/imz-managers/nis/about.html. Published 2018. Accessed January 23, 2019.

53. Agency for Healthcare Research and Quality. MEPS: Medical Expenditure Panel Survey. https://meps.ahrq.gov/mepsweb/. Published date unknown. Accessed January 23, 2020.

54. Child and Adolescent Health Measurement Initiative. About the National Survey of Children’s Health (NSCH). https://www.childhealthdata.org/learn-about-the-nsch/NSCH. Published date unknown. Accessed January 24, 2020.

55. National Center for Health Statistics. Data hosting: National Survey on Drug Use and Health (NSDUH). https://www.cdc.gov/rdc/b1datatype/nsduh.htm. Published 2018. Accessed January 23, 2019.

56. National Cancer Institute. HINTS: Health Information National Trends Survey. https://hints.cancer.gov/. Published date unknown. Accessed January 23, 2020.

57. National Institutes of Health, Food and Drug Administration. PATH: Population Assessment of Tobacco and Health. https://pathstudyinfo.nih.gov/UI/HomeMobile.aspx. Published 2018. Accessed January 20, 2020.

58. Lin CTJ, Zhang Y, Carlton ED, Lo SC. 2014 FDA Health and Diet Survey (HDS) topline report. https://www.fda.gov/media/96883/download. Published online 2016. Accessed January 30, 2020.

59. US Census Bureau. American Community Survey (ACS) data. https://www.census.gov/programs-surveys/acs/data.html. Published 2019. Accessed January 24, 2020.

60. US Census Bureau. About the Current Population Survey (CPS). https://www.census.gov/programs-surveys/cps/about.html. Published 2019. Accessed January 24, 2020.

61. US Census Bureau. American Housing Survey (AHS): About. https://www.census.gov/programs-surveys/ahs/about.html. Published 2020. Accessed January 24, 2020.

62. US Census Bureau. SIPP introduction & history. https://www.census.gov/programs-surveys/sipp/about/sipp-introduction-history.html. Published 2019. Accessed January 24, 2020.

63. US Bureau of Labor Statistics. American Time Use Survey (ATUS). https://www.bls.gov/tus/home.htm. Published date unknown. Accessed January 23, 2020.

64. US Bureau of Labor Statistics. Consumer Expenditure Surveys (CE). https://www.bls.gov/cex/. Publied date unknown. Accessed January 24, 2020.

65. National Center for Education Statistics. About the NHES. National Household Education Surveys Program (NHES). https://nces.ed.gov/nhes/about.asp. Published date unknown. Accessed December 20, 2019.

66. US Bureau of Labor Statistics. The NLSY79. National Longitudinal Surveys. https://www.bls.gov/nls/nlsy79.htm. Published 2020. Accessed April 20, 2020.

67. US Bureau of Labor Statistics. NLSY79 Children and Young Adults. National Longitudinal Surveys. https://www.bls.gov/nls/nlsy79-children.htm. Published 2020. Accessed April 20, 2020.

68. US Bureau of Labor Statistics. The NLSY97. National Longitudinal Surveys. https://www.bls.gov/nls/nlsy97.htm. Published 2020. Accessed April 20, 2020.

69. National Center for Education Statistics. Birth cohort (ECLS–B). https://nces.ed.gov/ecls/birthinstruments.asp. Published date unknown. Accessed August 8, 2019.

70. National Center for Education Statistics. Early Childhood Longitudinal Study (ECLS-K). In: NCES Handbook of Survey Methods. Washington, DC: National Center for Education Statistics; 2018.

71. National Center for Education Statistics. Early Childhood Longitudinal Study, Kindergarten Class of 2010–11 (ECLS-K:2011). In: NCES Handbook of Survey Methods. US Department of Education, Institute of Educational Sciences, National Center for Education Statistics; 2018.

72. National Center for Education Statistics. High School Longitudinal Study of 2009 (HSLS:09). https://nces.ed.gov/surveys/hsls09/index.asp. Published date unknown. Accessed February 6, 2020.

73. US Census Bureau, US Bureau of Justice Statistics. National Crime Victimization Survey (NCVS). https://www.census.gov/programs-surveys/ncvs.html. Published date unknown. Accessed January 24, 2020.

74. University of Michigan Institute for Social Research. Monitoring the Future: A Continuing Study of American Youth (MTF). http://monitoringthefuture.org/. Published 2019. Accessed January 24, 2020.

75. University of Michigan Institute for Social Research. HRS: The Health and Retirement Study. https://hrs.isr.umich.edu/about. Published 2019. Accessed January 24, 2020.

76. Standford University, University of Michigan. ANES: American National Election Studies. https://electionstudies.org/. Published 2020. Accessed February 5, 2020.

77. Smith TW. The General Social Surveys (GSS). Chicago, IL: NORC at the University of Chicago; 2016. (GSS Project Report No. 32)

78. NORC at the University of Chicago. National Survey on Teen Relationships and Intimate Violence (STRiV). https://www.norc.org/Research/Projects/Pages/survey-on-teen-realtionships-and-intimate-violence.aspx. Published date unknown. Accessed January 24, 2020.

79. Harvard University. Cooperative Congressional Election Study (CCES) frequently asked questions. https://cces.gov.harvard.edu/frequently-asked-questions. Published 2020. Accessed February 5, 2020.

80. Ramakrishnan K, Lee J, Lee T, Wong J. National Asian American Survey (NAAS). http://naasurvey.com/. Published date unknown. Accessed February 8, 2020.

81. Center for Studying Health System Change. CTS Household Survey and HSC Health Tracking Household Surveys. http://www.hschange.org/index8591.html. Published 2014. Accessed February 5, 2020.

82. Guttmacher Institute. 2012–2014 Continuity and Change in Contraceptive Use Study (CCCUS). https://www.guttmacher.org/population-center/dataset/2012-2014-continuity-and-change-contraceptive-use-study. Published 2020. Accessed February 5, 2020.

83. Alcohol Research Group. National Alcohol Surveys (NAS). http://arg.org/center/national-alcohol-surveys/. Published 2020. Accessed February 7, 2020.

84. American Psychological Association. Stress in America, United States, 2007–2018. RCMD: Resource Center for Minority Data. https://www.icpsr.umich.edu/icpsrweb/RCMD/studies/37288/summary. Published 2019. Accessed February 8, 2020.

85. University of Michigan Institute for Social Research. PSID: Panel Study of Income Dynamics. https://psidonline.isr.umich.edu/. Published 2020. Accessed January 24, 2020.

86. US Census Bureau. Household Pulse Survey: Measuring household experiences during the coronavirus pandemic. https://www.census.gov/data/experimental-data-products/household-pulse-survey.html. Published December 22, 2021. Accessed December 29, 2020.

87. National Endowment for the Arts. Arts Basic Survey, United States, 2020 (ICPSR 37972). Ann Arbor, MI: Inter-university Consortium for Political and Social Research [distributor]; 2021. (doi: 10.3886/ICPSR37972.v1).

88. Centers for Disease Control and Prevention. The National Intimate Partner and Sexual Violence Survey (NISVS). https://www.cdc.gov/violenceprevention/datasources/nisvs/index.html. Published June 4, 2020. Accessed March 7, 2020.

89. US Census Bureau. National Longitudinal Mortality Study (NLMS). https://www.census.gov/nlms. Published 2019. Accessed January 24, 2020.

90. Board of Governors of the Federal Reserve System. Survey of Household Economics and Decisionmaking (SHED). https://www.federalreserve.gov/consumerscommunities/shed.htm. Published May 17, 2021. Accessed December 30, 2021.

91. Pew Charitable Trusts. *Survey of American Family Finances, [United States]*, *2014-2015.* Ann Arbor, MI: Inter-university Consortium for Political and Social Research [distributor]; 2020. (doi:10.3886/ICPSR37629.v2)

92. NORC at the University of Chicago. National Survey of Early Care and Education (NSECE). https://www.norc.org/Research/Projects/Pages/national-survey-of-early-care-and-education.aspx. Published date unknown. Accessed January 12, 2022.

93. National Center for Health Statistics. About RANDS. https://www.cdc.gov/nchs/rands/about.htm. Published June 8, 2020. Accessed January 17, 2022.

94. National Center for Health Statistics. Birth data. https://www.cdc.gov/nchs/nvss/births.htm. Published 2019. Accessed January 24, 2020.

95. National Center for Health Statistics. Linked birth and infant death data. https://www.cdc.gov/nchs/nvss/linked-birth.htm. Published 2018. Accessed January 23, 2020.

96. 96. National Center for Health Statistics. Fetal deaths. https://www.cdc.gov/nchs/nvss/fetal_death.htm#Methods. Published 2018. Accessed February 8, 2020.

97. National Center for Health Statistics. Mortality data. https://www.cdc.gov/nchs/nvss/deaths.htm. Published 2019. Accessed January 23, 2020.

98. University of Michigan. National Poll on Healthy Aging (NPHA). https://www.healthyagingpoll.org/. Published 2021. Accessed March 23, 2021.

99. Stewart A. Changes in Federal Surveys Due to and during COVID-19. Minneapolis, MN: State Health Access Data Assistance Center (SHADAC); 2021.

100. US Census Bureau. Impact of Pandemic on the American Community Survey. https://www.census.gov/newsroom/press-kits/2021/impact-pandemic-2020-acs-1-year.html. Published July 29, 2021. Accessed October 30, 2021.

101. Rothbaum J, Bee A. Coronavirus Infects Surveys, too: Survey Nonresponse Bias and the Coronavirus Pandemic. Washington, DC: United States Census Bureau; 2021.

102. US Bureau of Labor Statistics. Effects of COVID-19 pandemic and response on the Consumer Expenditure Surveys. https://www.bls.gov/covid19/effects-of-covid-19-pandemic-and-response-on-the-consumer-expenditure-surveys.htm. Published September 9, 2021. Accessed October 30, 2021.

103. US Census Bureau. Research Data Centers. https://www.census.gov/about/adrm/fsrdc/locations.html. Published November 18, 2021. Accessed January 23, 2022.

104. Li RM, McCardle P, Clark RL, et al. Diverse Voices: The Inclusion of Language-Minority Populations in National Studies: Challenges and Opportunities. Bethesda, MD: National Institute on Aging; 2001.

105. Brown JD, Heggeness ML, Dorinski SM, et al. Predicting the effect of adding a citizenship question to the 2020 Census. Demography. 2019;56(4):1173–1194. (doi:10.1007/s13524-019-00803-4).

106. National Center for Health Statistics. NHIS - Survey Reports and Data Linked to NHIS. Published October 21, 2020. Accessed January 30, 2021.

107. US Census Bureau. Census Longitudinal Infrastructure Project (CLIP). https://www.cdc.gov/nchs/nhis/nhis_products.htm. Published date unknown. Accessed January 26, 2021.

108. Wang HL. Biden administration tables Trump’s citizenship data request for redistricting. https://www.npr.org/2021/01/22/959609263/biden-administration-tables-trumps-citizenship-data-request-for-redistricting. Published January 22, 2021. Accessed January 27, 2021.

