## Appendix A Data extraction form for "A Systematic Review of Datasets to Research the Effect of Policy and Place on Immigrants’ Health"

Citation:

### **Study overview**

Years available:

Study description:

Target population:

Type of study:

- Cross-sectional
- Longitudinal
  - Cohort
  - Panel

Survey mode: Check all that apply. Indicate the main survey mode, if applicable.

- In-person
- Telephone
- Online
- On paper (e.g., by mail, classroom questionnaires)

Sample size in most recent year:

Oversamples:

Domains of health:

- Health care access
  - Health insurance coverage
  - Health care utilization (e.g., check-ups, unmet needs)
- Self-rated health or parent-rated general health
- Physical health
- Mental or behavioral health
- Disability / health-related limitations
- Health behaviors
- Other (specify): ­­_____________________________________________________

### **Geography**

Geographic coverage:

Public-use geocodes:

- State
- MSA / CBSA
- County
- Census Place
- Zip code
- Census tract
- Census block group
- Census block
- Latitude / longitude

Restricted-use geocodes:

- State
- MSA / CBSA
- County
- Census Place
- Zip code
- Census tract
- Census block group
- Census block
- Latitude / longitude

Geography notes (e.g., are some cities / counties always included?):

### **Race and ethnicity**

Public-use race categories:

- White
- Black
- American Indian or Alaska Native (combined)
- American Indian
- Alaska Native
- Asian, Native Hawaiian, or other Pacific Islander (combined)
- Asian
  - Asian Indian
  - Chinese
  - Filipino
  - Japanese
  - Korean
  - Vietnamese
  - Other Asian (specify): ­­__________________________________________
- Native Hawaiian or other Pacific Islander (combined)
  - Native Hawaiian
  - Guamanian or Chamorrow
  - Samoan
  - other Pacific Islander (specify): ­­_______________________________________
- Other race (combined category)
- Multiple race (combined category)
- Other race groups identifiable (specify): _____________________________________ ______________________________________________________________ _____________________________________

Restricted-use race categories:

- White
- Black
- American Indian or Alaska Native (combined)
- American Indian
- Alaska Native
- Asian, Native Hawaiian, or other Pacific Islander (combined)
- Asian
  - Asian Indian
  - Chinese
  - Filipino
  - Japanese
  - Korean
  - Vietnamese
  - Other Asian (specify): ­­__________________________________________
- Native Hawaiian or other Pacific Islander (combined)
  - Native Hawaiian
  - Guamanian or Chamorrow
  - Samoan
  - other Pacific Islander (specify): ­­_______________________________________
- Other race (combined category)
- Multiple race (combined category)
- Other race groups identifiable (specify): _____________________________________ ______________________________________________________________ _____________________________________

For multiple race individuals, can researchers identify specific racial groups (e.g., White + Black, Black + AIAN)?

Public-use Hispanic/Latino origin groups:

- Mexican
- Puerto Rican
- Cuban
- Central or South American (combined)
- Central American
- South American
- Dominican
- Salvadoran
- Spanish (from Spain)
- Other Hispanic/Latino groups (specify): _________________________________ ______________________________________________________________ _____________________________________

Restricted-use Hispanic/Latino origin groups:

- Mexican
- Puerto Rican
- Cuban
- Central or South American (combined)
- Central American
- South American
- Dominican
- Salvadoran
- Spanish (from Spain)
- Other Hispanic/Latino groups (specify): _________________________________ ______________________________________________________________ _____________________________________

For multiple Hispanic origin individuals, can researchers identify specific groups (e.g., Mexican + Puerto Rican, Mexican + Salvadoran)?

Is Hispanic ethnicity asked separately from race?

### **Immigration**

Which of the following are available in public-use data?

- Nativity: Identify US-born vs. foreign-born
- Nativity: Identify specific country of birth
- Nativity: Identify parent(s) as US-born vs. foreign-born
- Nativity: Identify parent(s)’ specific country of birth
- Migration: Year arrived in US, age arrived in US, or years living in US, in categories
- Migration: Year arrived in US, age arrived in US, or years living in US, continuous
- Legal status: Citizenship
- Legal status: Identify legal status for noncitizens (specify which legal statuses can be identified: ­­­___________________________________________________________ ____________________________________________________________________)
- Other migration-related variables (specify: ­­__________________________________ ____________________________________________________________________________________________)

Which of the following are available in restricted-use data?

- Nativity: Identify US-born vs. foreign-born
- Nativity: Identify specific country of birth
- Nativity: Identify parent(s) as US-born vs. foreign-born
- Nativity: Identify parent(s)’ specific country of birth
- Migration: Year arrived in US, age arrived in US, or years living in US, in categories
- Migration: Year arrived in US, age arrived in US, or years living in US, continuous
- Legal status: Citizenship
- Legal status: Identify legal status for noncitizens (specify which legal statuses can be identified: ­­­___________________________________________________________ ____________________________________________________________________)
- Other migration-related variables (specify: ­­__________________________________ ____________________________________________________________________________________________)

### **Language**

Languages in which interviews are conducted:

Language variables in public-use data:

- Language of interview
- English proficiency
- Language(s) usually spoken in the home
- Other language variables (specify: ­­____________________________________________ ____________________________________________________________________________________________)

Language variables in restricted-use data:

- Language of interview
- English proficiency
- Language(s) usually spoken in the home
- Other language variables (specify: ­­____________________________________________ ____________________________________________________________________________________________)

### **Availability of restricted data**

Apply through:

Timeline:

Requires RDC access?

Fees:
